# Optimal COVID-19 quarantine and testing strategies

**DOI:** 10.1101/2020.10.27.20211631

**Authors:** Chad R. Wells, Jeffrey P. Townsend, Abhishek Pandey, Seyed M. Moghadas, Gary Krieger, Burton Singer, Robert H. McDonald, Meagan C. Fitzpatrick, Alison P. Galvani

## Abstract

As economic woes of the COVID-19 pandemic deepen, strategies are being formulated to avoid the need for prolonged stay-at-home orders, while implementing risk-based quarantine, testing, contact tracing and surveillance protocols. Given limited resources and the significant economic, public health, and operational challenges of the current 14-day quarantine recommendation, it is vital to understand if shorter but equally effective quarantine and testing strategies can be deployed. To quantify the probability of post-quarantine transmission upon isolation of a positive test, we developed a mathematical model in which we varied quarantine duration and the timing of molecular tests for three scenarios of entry into quarantine. Specifically, we consider travel quarantine, quarantine of traced contacts with an unknown time if infection, and quarantine of cases with a known time of exposure. With a one-day delay between test and result, we found that testing on exit (or entry and exit) can reduce the duration of a 14-day quarantine by 50%, while testing on entry shortened quarantine by at most one day. Testing on exit more effectively reduces post-quarantine transmission than testing upon entry. Furthermore, we identified the optimal testing date within quarantines of varying duration, finding that testing on exit was most effective for quarantines lasting up to seven days. As a real-world validation of these principles, we analyzed the results of 4,040 SARS CoV-2 RT-PCR tests administered to offshore oil rig employees. Among the 47 positives obtained with a testing on entry and exit strategy, 16 cases that previously tested negative at entry were identified, with no further cases detected among employees following quarantine exit. Moreover, this strategy successfully prevented an expected nine offshore transmission events stemming from cases who had tested negative on the entry test, each one a serious concern for initiating rapid spread and a disabling outbreak in the close quarters of an offshore rig. This successful outcome highlights that appropriately timed testing can make shorter quarantines more effective, thereby minimizing economic impacts, disruptions to operational integrity, and COVID-related public health risks.

## Introduction

The COVID-19 pandemic has engendered unprecedented efforts to quell ongoing outbreaks and manage healthcare capacity, including strict travel restrictions and stay-at-home orders. These efforts have disrupted workplaces, leading to significant and pervasive socioeconomic costs ^1,2^. In turn, these economic pressures have led many governments and corporations to lift restrictions^3^. Safely reopening in the absence of a vaccine relies on reducing the likelihood of an infectious individual entering a workplace, school, or other social gathering ^4^. Current strategies to ensure safety often include a 14-day quarantine—either as a consequence of travel or following exposure to an infected person, as recommended by the World Health Organization (WHO).^5^ These quarantines are sometimes combined with entry and/or exit testing, in which a positive test prompts isolation until recovery.

Quarantine imposes myriad challenges for institutions of government, militaries, businesses, universities, and other entities. At the individual level, the recommended 14-day quarantine causes strain on mental health. ^6,7^ This burden is coupled with the associated economic toll and potential impacts on operational integrity. For example, the typical 14-day on-and-off cycle for offshore oil and gas employees is substantially disrupted when quarantine measures are required. These quarantines result in prolonged time periods that crew members are away from their home. Given the impact of long quarantines on mental health ^6,7^, we evaluated the potential that a shorter quarantine combined with testing optimization could achieve reduced transmission of COVID-19 within close-quarter environments where there is potentially a high risk for rapid spread.

Evidence suggests that isolation of cases upon symptom onset is insufficient to contain an outbreak of COVID-19 ^8^. The likelihood of transmission can be reduced substantially through quarantine and testing ^4^. Previous work has focused on the impact of quarantine and testing on population-level COVID-19 incidence and deaths ^9–11^, shortened quarantines upon negative RT-PCR test at entry from contact tracing or seven days after exposure ^12^ and testing measures that are most appropriate for disease surveillance within a high-risk population (e.g. healthcare workers) by examining various testing frequencies and their reduction of secondary infections^13^. Currently, there is no consensus regarding the optimal duration of quarantine or timing of testing that minimizes the probability of post-quarantine transmission (PQT), defined as one or more infections observed after the quarantine period. Many institutions are relying on testing at entry into quarantine combined with other measures such as symptom screenings, hand sanitizers, and face masks to reduce the risk of an outbreak. However, the majority of COVID-19 transmission is attributable to pre-symptomatic and asymptomatic cases screening for symptoms alone is inadequate to prevent or interrupt a COVID-19 outbreak ^8^. In addition, testing too early post-infection is likely to produce a false-negative result ^14^. Thus, symptom-based screening and one-time testing could still entail a significant probability of PQT.

Some jurisdictions have suggested and implemented testing upon exit from a 14-day quarantine^15^. For example, Australia has implemented a compulsory 14-day quarantine, with testing within 48 hours after arrival and between day 10 and 12 of quarantine, to reduce transmission from imported cases ^16^. Although these multiple tests aid in case identification, this strategy does not include any reduction of the burden of long quarantine. Understanding the complementarity of quarantine and testing in reducing PQT would provide vital insight into effective strategies that mitigate disease spread in travel-based and contact-tracing based contexts.

We applied a mathematical modeling approach to evaluate whether a less burdensome quarantine and testing strategy exists that would be epidemiologically equivalent to the standard 14-day quarantine protocol in reducing PQT. This model accounts for the infectivity profile of an infected individual as well as the temporal diagnostic sensitivity of RT-PCR testing. Across a variety of quarantine and testing scenarios, we estimated the probability of PQT for an infected individual who has not manifested symptoms by the end of the quarantine period. We considered three applications: (i) quarantine for travel, initiated at random times across the infectious course, (ii) quarantine prompted by contact-tracing and therefore initiated early in the infectious course, and (iii) quarantine when the time of exposure is known. We compared the probability of PQT under three testing scenarios: (i) on entry to quarantine only, (ii) on exit from quarantine only, and (iii) on both entry to and exit from quarantine for an infected individual. Across these scenarios, we varied the duration of quarantine and identified the optimal testing date based on that duration. As validation of our recommendations, we analyzed the real-world application of our model-based findings to protocols within the oil and gas industry that prevented offshore transmission.

## Results

We derived an infectivity profile based on transmission pairs of COVID-19 infected individuals ^17^, a basic reproduction number of *R*_0_ = 2.5, and an incubation period of 8.29 days ^18^, and estimated the temporal diagnostic sensitivity of RT-PCR tests ^19^ (**Table S1**). Specifying 30.8% of infections as remaining asymptomatic across the disease course ^20,21^, we estimated that perfect isolation of cases upon symptom onset would reduce the reproduction number to 1.6, with 1.2 secondary cases occurring during the incubation period (**Fig. S1A**). The reproductive number remained above one even when we lowered the asymptomatic proportion to 22.6% or reduced *R*_0_ to 2 (**Fig. S1B–D**). Therefore, perfect isolation of all symptomatic individuals would not be sufficient to interrupt the chain of disease transmission.

### Entry into quarantine when the time of exposure is unknown

For settings where there is no administrative knowledge of the time of exposure such as travel quarantine, we computed the expected PQT (**Fig. S2**) and the probability of PQT after a range of quarantine durations without testing (**Fig. 1A, Fig. S3A**). Assuming individuals self-isolate immediately upon symptom onset, the probability of PQT declines as the duration of quarantine increases (**Fig. 1A**). This probability is less than 0.25 with a quarantine duration of at least three days, and falls below 0.05 for quarantines of eight days or longer.

**Figure 1:**
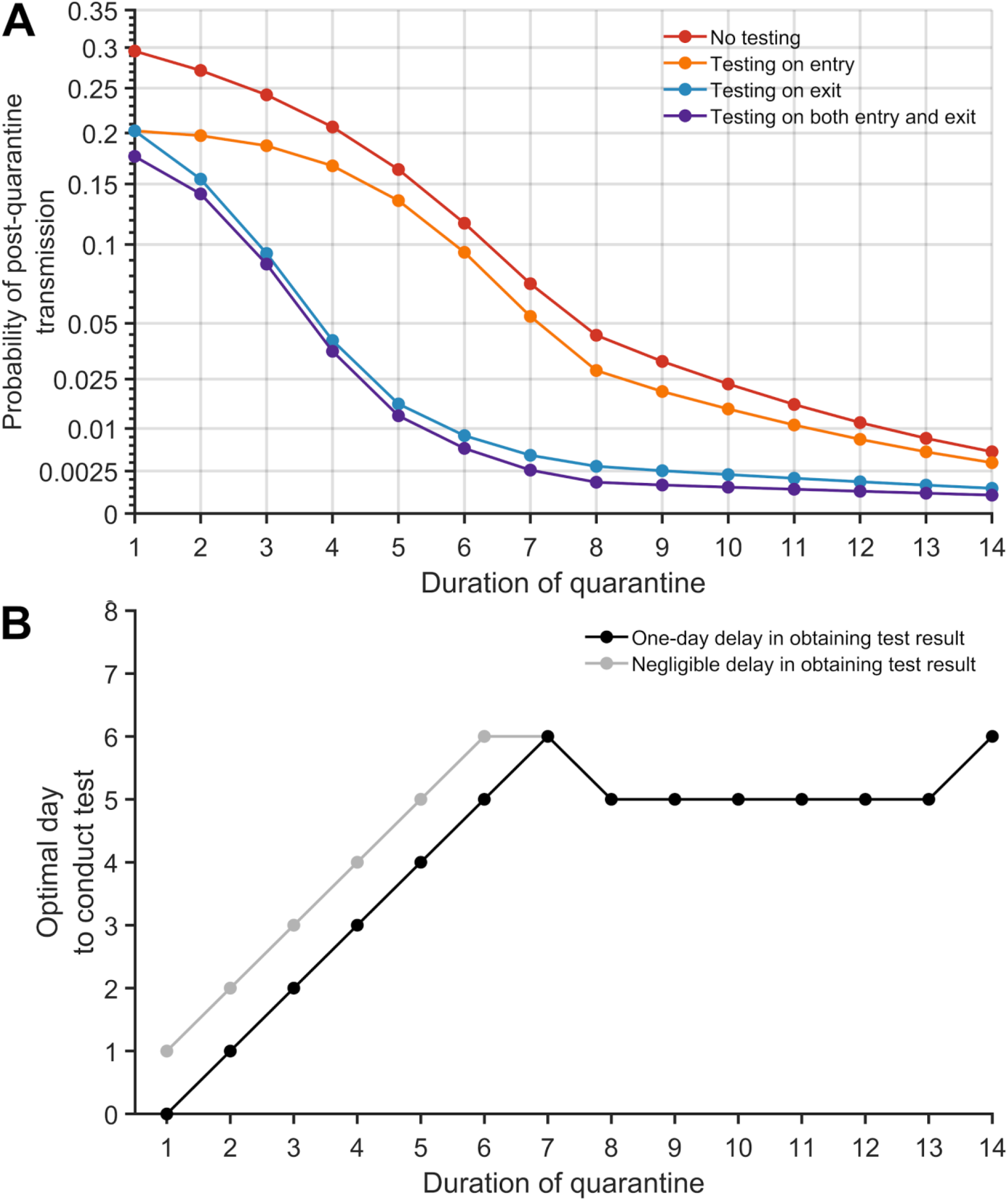
The probability of post-quarantine transmission and optimal day to conduct test when an infected individual enters quarantine uniformly within the incubation or asymptomatic period, for no testing and three testing strategies, and durations of quarantine from 1–14 days, with an incubation period of 8.29 days, 30.8% asymptomatic infections and perfect self-isolation of symptomatic infections. **(A)** Curves for the probability of post-quarantine transmission (one or more post-quarantine infections) without testing (red), with testing upon entry to quarantine (orange), on exit from quarantine (blue), and on both entry to and exit from quarantine (purple). Results include a one-day delay in sample collection to results, such that testing on exit occurred the day before the end of quarantine. (**B**) The optimal day to test during quarantine with a one-day delay (black) and a negligible delay (gray) in obtaining test results.

The impact of quarantine can be augmented through testing. We assumed a 24-hour delay between the sample collection and test results, so that testing on exit occurred one day before the end of quarantine. Individuals who tested positive or developed symptoms were isolated until recovery. We found that any testing during quarantine contributed to a reduction in the probability of PQT across the full range of quarantine duration (**Fig. 1A** and **Fig. S3A)**. The magnitude of this reduction was dependent on both the duration of quarantine and the timing of the testing.

The largest reduction in the probability of PQT from conducting a single test occurred when it was performed on exit for quarantines of seven days or less; on day five for quarantines lasting between eight and 13 days; and on day six for quarantines that are 14 days or longer (**Fig. 1B**). As quarantined (asymptomatic) cases proceed through their quarantine, they simultaneously progress through their infectious course. Symptom onset will send a substantial fraction of infected individuals to isolation and diagnostic sensitivity decreases for the remainder^19^, leading to slightly diminishing benefits of “exit” tests performed later than day six.

Comparing the three testing strategies, we found that testing on both entry and exit from quarantine provides the greatest reduction in PQT, whereas the benefit of testing at entry is minimal (**Fig. 1A, Fig. S3A**). Testing on exit consistently and substantially outperformed testing on entry across all quarantine durations considered (**Fig. 1A**).

We specifically compared strategies of quarantine and testing against the widely implemented WHO recommendation to quarantine for 14 days (without testing) ^5^. In this comparison, a 13-day quarantine with testing on entry, a seven-day quarantine with testing on exit, and a seven-day quarantine with testing on both entry and exit each provide equivalent or lower probabilities of PQT (**Fig. 1A, Fig. S3**).

### Assessment of quarantine and testing strategies implemented for offshore facilities

We applied our results in the context of employees of an off-shore oil company who were working a cycle of 26 days on, then 16 days off, a schedule that had been modified to make efficient use of a mandatory quarantine that was implemented during the pandemic. During the early stages of the epidemic, when prevalence was low, a three-day quarantine had been implemented by the company in a secured facility, with testing on entry. Our risk-reduction models indicated substantial marginal benefit for increasing quarantine to 5–7 days with a test on exit. Testing on entry was retained for operational purposes, and testing 96 h later was initiated, accompanied by expansion to a seven-day quarantine for Region A and a five-day quarantine for Region B.

To assess the practical implications of our recommendations, 4040 RT-PCR tests were conducted in region A and region B (serviced by different laboratories) prior to travel to offshore rigs. Among these, 69 results were positive (1.7%). Of the 1792 RT-PCR tests conducted as tests on entry when the initial three-day quarantine was in effect, there were 22 positive results (1.2%). After advisement, Region A deployed a seven-day home quarantine for all cycles starting August 13, where testing was performed on entry and exit (96 h after the first test); 50.0% (1/2) of the positive tests occurred on exit, following a negative test on entry (**Fig. 2A**). Starting June 25, Region B expanded to a five-day hotel quarantine with testing on both entry and 96 h after the first test. For the period in which this strategy was implemented, 33.3% (15/45) of the positive tests were obtained upon the exit test, following a negative entry test (**Fig. 2B**). Further validation of the entry and exit testing protocol was provided through an additional 155 RT-PCR tests performed post-quarantine (11 days after the initial test) in Region B, all of which were negative.

**Figure 2:**
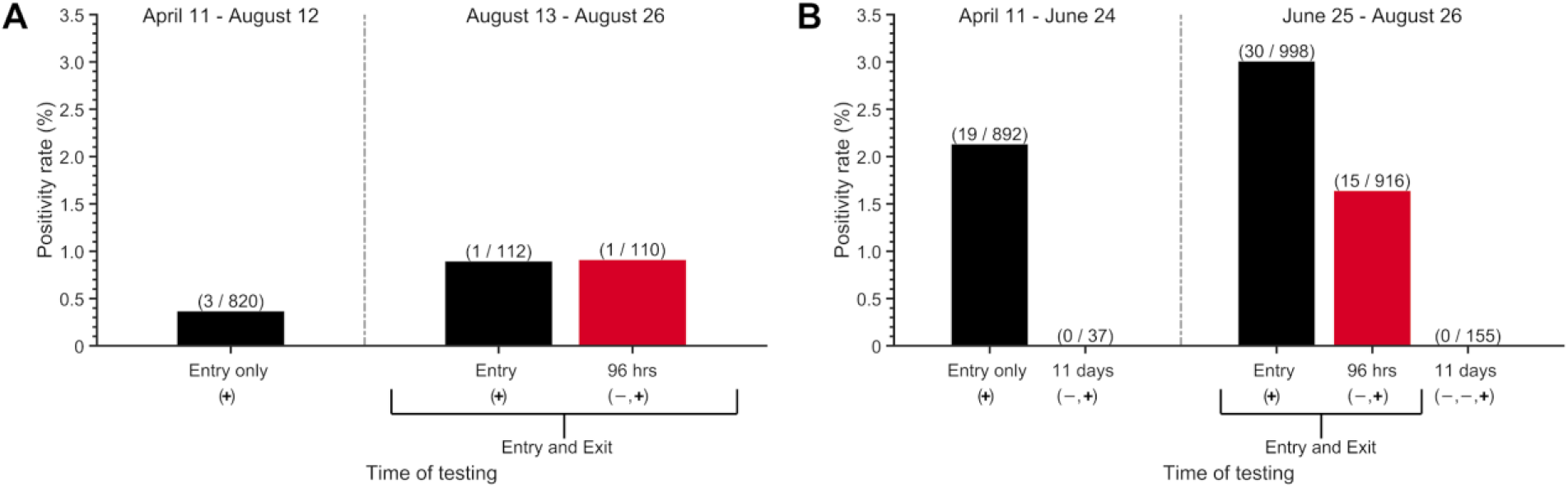
Weekly SARS-CoV-2 testing and positivity rate between April 11 to August 26, 2020, within two regions where crew members were quarantined: (**A**) region A, with a seven-day quarantine, where testing on entry and exit was started on August 13, and (**B**) region B, with a five-day quarantine, where testing on entry and exit was started on June 25. Initially, a three-day quarantine with testing only on entry was conducted in both regions. The vertical dashed line separates the early strategy of testing on only entry (left) and the later strategy of testing on both entry and exit (right), including follow-up post-quarantine tests conducted 11 days after the initial test (i.e., on day 12). Negative and positive sequential symbols − and + indicate the test histories. In these results, negative symbols are always conveying results to tests that were previous to the results quantified by the bar above. The number of positive tests (numerator) and the number of tests conducted (denominator) is denoted above the bar in parentheses.

No offshore worker registering negative tests on entry and on exit from quarantine was later diagnosed with COVID-19 during their offshore work. To quantify the added benefit of the test at 96 h, we calculated the probability of PQT for the cases detected by this second test. Compared with a three-day quarantine and testing only on entry, extending the quarantine duration and adding testing on exit (96 h after the first test) reduced the probability of PQT by 98% for the seven-day quarantine and 93% for a five-day quarantine. If the single case identified on the exit test from region A had remained undetected within the seven-day quarantine, we estimate an off-shore probability of PQT of 0.13. If the 15 cases that had been ascertained on exit from region B had remained undetected after the five-day quarantine without testing on exit, we estimate that the probability of at least one event of PQT would have been 0.99, and would have resulted in an expected 9 offshore transmission events—each one a serious concern for initiating further rapid spread and a disabling outbreak in the close quarters of an offshore rig.

### Accounting for prevalence of disease in the community

We evaluated the impact of disease prevalence in the community on the probability of PQT (**Fig. S6**). For a cohort of 40 individuals undergoing a five-day quarantine with prevalence of 1%, we estimated the probability of PQT to be 0.06 for testing only on entry, and 0.005 for testing on both entry and exit (**Fig. S6B**). For a seven-day quarantine and the same prevalence, the probability of PQT drops from 0.02 for testing only on entry to 0.001 when augmented with testing on exit (**Fig. S6C**).

### Contrasting contact tracing and uniform entry into quarantine

Contact tracing is ideally initiated following identification of a positive case either by symptom presentation or by surveillance screening through testing. We evaluated the impact of quarantine initiated through contact tracing on reducing PQT under scenarios of no delay (**Fig. 3A, Fig. S7–S8**) or one-day delay in outreach to exposed contacts (**Fig. S9–S10**). Tracing of contacts was assumed to be initiated by the onset of relevant COVID-19 symptoms. Rapid contact tracing results in the quarantine of infected contacts early in their infection course, thereby increasing the recommended duration of quarantine and changing the relationship between test timing and the probability of PQT, compared to uniform entry into quarantine (**Fig. 3A** vs **Fig. 1A)**.

**Figure 3:**
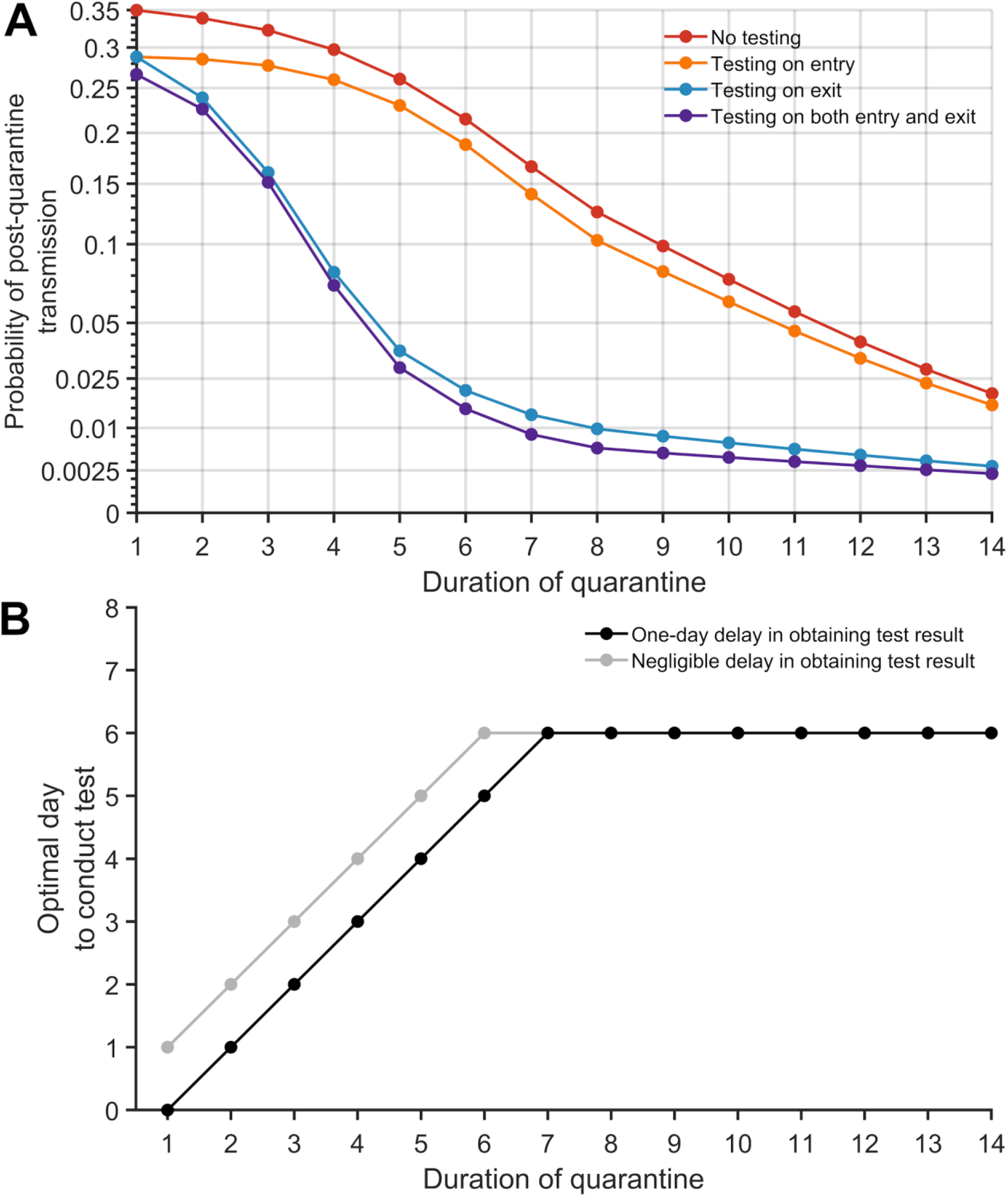
The probability of post-quarantine transmission for no testing and three testing strategies applied to 1–14-day durations of quarantine, when an individual enters quarantine through contact tracing, specifying an incubation period of 8.29 days, 30.8% asymptomatic infections, and perfect self-isolation of symptomatic infections. (**A**) The probability of one or more post-quarantine infections without testing (red), with testing upon entry to quarantine (orange), on exit from quarantine (blue), and on both entry to and exit from quarantine (purple), assuming that testing on exit occurs on the penultimate day of quarantine. (**B**) The optimal day to test during quarantine for a specified quarantine duration, with that one-day delay (black) and with a negligible delay (gray) in obtaining test results.

However, the combination of shorter quarantines with exit testing maintains high effectiveness compared with 14-day quarantines without testing. When cases are identified through contact tracing, we found that a seven-day quarantine with testing on exit and a six-day quarantine with testing on entry and exit each result in an probability of PQT equivalent or lower than a 14-day quarantine with no testing; testing on entry bestowed only trivial benefit (**Fig. 3A, Fig. S8**). For quarantines of seven days or less, the optimal test timing was upon exit. For quarantines beyond seven days, the optimal timing was day six (**Fig. 3B**).

### Optimal day of testing for a known time of exposure

When a specific date of exposure can be identified for a traced contact, the optimal test timing differs from that calculated by integrating over all possible exposure times. When quarantined one day post-infection and tested on entry, an additional test on day six of quarantine is optimal; the optimal day of testing then decreases linearly. For an individual entering quarantine seven or more days post-infection, the optimal test date is the test on entry (**Fig. 4**).

**Figure 4:**
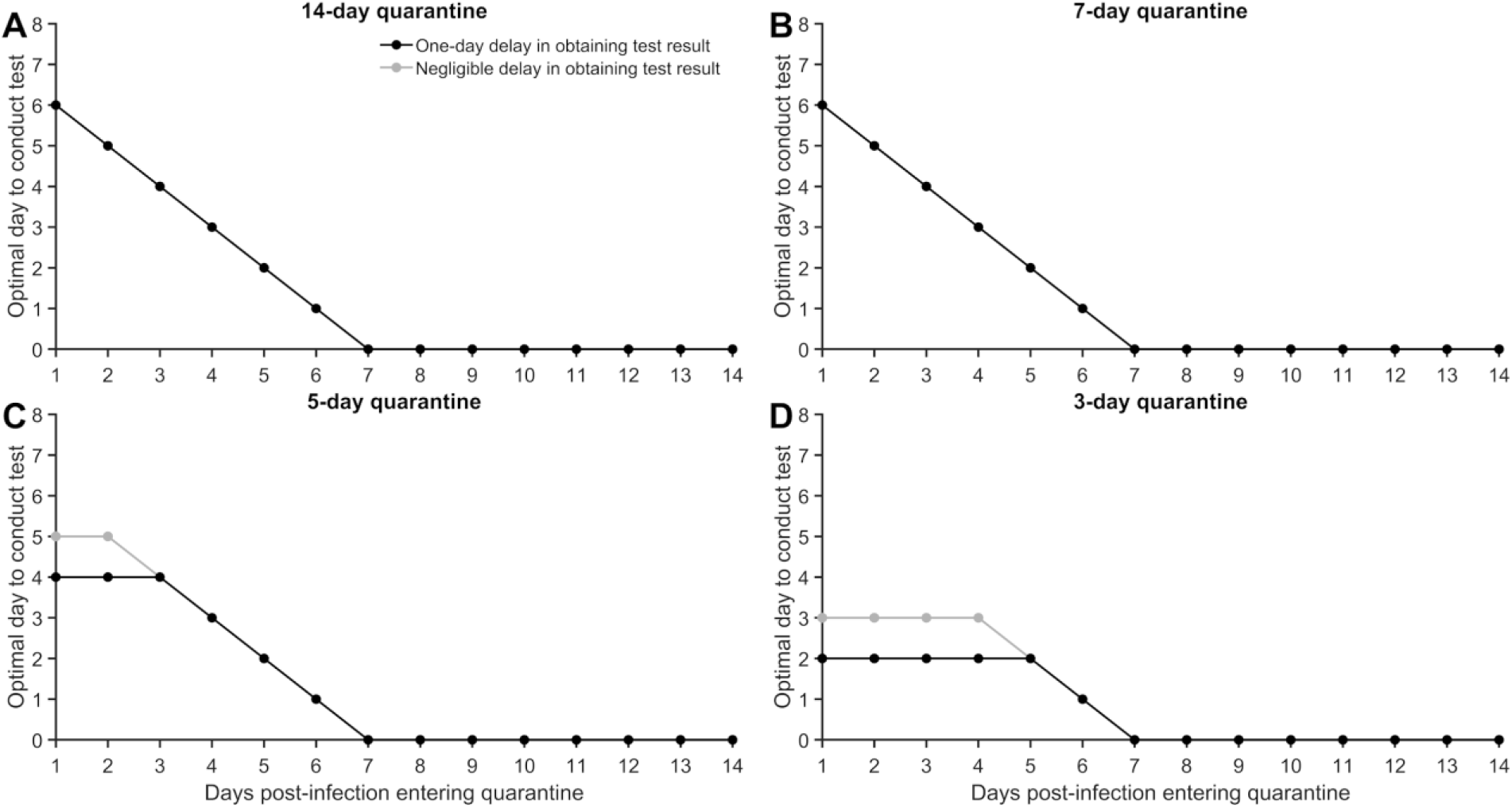
For a case whose date of exposure has been identified as occurring 1–14 days prior to quarantine, the optimal day to conduct the RT-PCR test, assuming perfect self-isolation of symptomatic infections, 30.8% asymptomatic infections, an incubation period of 8.29 days, and a quarantine lasting (**A**) 14 days, (**B**) seven days, (**C**) five days, and (**D**) three days.

### Sensitivity analyses

We performed a comparative analysis specifying a latent period that is one day greater or lesser than the reported 2.9 days ^22^. The expected number of secondary cases occurring before symptom onset was similar among the different latent periods (1.21 infection for a latent period 2.9 days; 1.24 infections for a latent period of 1.9 days; and 1.27 infections for a latent period of 3.9 days). The infectivity profiles differed among the three latent periods, with a peak infectivity that is higher for both the 1.9-day and 3.9-day latent periods when compared to our baseline (**Fig. S11**).

For quarantine periods of at least seven days and individuals entering quarantine uniformly across the time course of infection (Fig. S12–S15), the probability of PQT was lower for shorter latent periods. For shorter quarantines, the relationship between the probability of PQT and latent period is more intricate. For traced contacts entering quarantines of eight days or longer as (**Fig. S16–S19**), shorter latent periods entailed lower probability of PQT. For traced contacts entering quarantines of fewer than eight days, the relationship of latent period to probability of PQT is more complex. However, one-day changes in the latent period affect the optimal day to conduct a single test by at most one day (**Fig. S4)**. Specifically, we found that a 3.9-day latent period decreased the optimal day of testing estimated for a 2.9-day latent period, whereas a 1.9-day latent period increased the best day to conduct a single test.

For our analysis of potential outbreaks consequent to offshore-rig quarantine and testing, we analysed sensitivity of our result to the proportion of asymptomatic individuals on the probability of PQT (**Fig. S5**). We found that the estimated probability of PQT using the strategy of testing upon entry and at 96 h moderately increased if a higher proportion of infections were expected to be asymptomatic (**Fig. S5**).

## Discussion

Here, we derived theory to calculate the probability of post-quarantine transmission of COVID-19 for a wide range of durations of quarantine, supplemented by testing on entry to quarantine, on exit from quarantine, or both. For quarantines with durations of up to seven days, we found that testing on exit provided the greatest marginal benefit in terms of reducing the probability of PQT. Testing on entry provided modest benefits in combination with quarantine or with testing on exit. For a quarantine with a duration longer than seven days, the optimal testing time is on day five or six. Optimal testing times were fairly consistent between travel quarantines and quarantines of traced contacts, differing at most by a day. The benefits of testing later in quarantine were demonstrated by test results of oil crewmembers heading offshore that identified 16 cases testing negative on entry and positive on exit that could easily have resulted in costly and logistically difficult-to-handle offshore outbreaks. When the time of exposure is known, the optimal day for a test for quarantines of a week or more starts at day six of the quarantine, decreasing linearly to day-of-entry for individuals who have been infected for seven or more days. It may seem counter-intuitive that the optimal test for so many identified timings of exposure is on entry, yet testing on entry has so much less impact than testing on exit when the date of exposure is unknown. Indeed, for individuals that are tested after the incubation period (e.g. later than symptom onset), the diagnostic sensitivity of the RT-PCR test has started to decline. However, for individuals late in disease, there is also far less infectivity left in their disease course. The high remaining infectivity of individuals early in disease course markedly outweighs the low infectivity of individuals late in disease course in influencing the optimal day of testing to prevent post-quarantine transmission.

An outbreak can be triggered or sustained within an environment that is monitored only for symptoms of COVID-19. Quarantining individuals before returning to work or school has been a common strategy among many businesses, the military and universities to prevent potential outbreaks ^23,24^. An offshore or military setting is one of numerous close-quarters environments in modern society where an outbreak can seriously impact operational integrity, leading to compromised safety and adverse economic consequences. Hence, minimizing outbreak risk while maintaining staffing is critical. Testing may allow for the quarantine duration to be reduced without increasing the risk of PQT. For example, many universities have implemented plans for quarantining and frequent testing of students and employees, where resources allow ^25,26^. For businesses and close-quarters environments, the impact of false negatives is a substantially greater issue for operational integrity than false positives. Consistent with the results from our analytic model (**Fig. 1A** and **Fig. 3A**), simulations from a recent agent-based model suggest that testing on exit—or entry and exit—of a seven-day quarantine can avert similar transmission as a 14-day quarantine with no transmission ^12^. Our results show that testing upon entry to quarantine carries such a risk of false negatives, as infected individuals who enter quarantine very early in the incubation period of disease may not be detected due to low viral loads.

Our estimates for the probability of PQT for the various strategies were estimated assuming a basic reproductive number of 2.5 throughout the disease course, and unchanged post-quarantine. In the offshore environment, individuals are living in very confined quarters which could lead to higher post-quarantine transmission and a larger number of secondary infections. In some community settings, the number of secondary infections can be reduced through mask-wearing, social distancing, and other non-pharmaceutical interventions. These changes in the number of secondary infections post-quarantine can markedly influence the probability of PQT. However, they would not affect the relative benefit of testing on exit compared to entry. Therefore, our qualitative finding of the optimality of testing later in quarantine than on entry are robust to settings with extensive post-quarantine transmission.

As prevalence in the general community increases (**Fig. S6**, blue and purple), there are benefits to conducting additional tests during quarantine: as substantial numbers of infected individuals enter quarantine, larger numbers of individuals may proceed through testing with rare false-negative test results, increasing PQT. Addressing false negatives that inevitably occur at high prevalence can be aided by performing additional tests during quarantine; the impact of any specific set of tests can be quantified within our model framework. In future research, the theory can be applied to evaluate the impact of incorporating recent innovations such as saliva RT-PCR tests and rapid antigen tests. These alternate approaches could exhibit altered optima. We have not quantified more extensive testing strategies here due to the limited availability of testing, potentially high and largely unknown correlations among false-negative test results for individual cases, and the observed moderate marginal benefit of additional testing performed in early stages of disease with lower detection rates (**Fig. S28**).

Optimal timing of limited testing during quarantine improves the ability to control PQT. Testing several days into quarantine increases the likelihood of an infected case testing positive (**Fig. S4**). The increasing diagnostic sensitivity of the RT-PCR test is attributable to the rapidly increasing viral load following the less detectable latent stage of infection. If the infected individual remains asymptomatic, testing near the end of a standard 14-day quarantine can also lead to low diagnostic sensitivity due to a declining viral load as they overcome the infection ^27^. Australia has implemented a mandatory 14-day quarantine for individuals arriving into the country, with testing during the first two days of arrival and between day 10 and 12 of quarantine ^16^. Though the differences are moderate, our analysis indicates that the lowest probability of PQT is achieved by testing on day six of the standard 14-day quarantine (**Fig. 1B, Fig 3B**).

Testing was found to result in a smaller reduction of the expected PQT when cases enter quarantine through contact tracing compared to when they enter as a consequence of travel regulation. Contact tracing will usually identify more infected cases per quarantined individual than will travel quarantine, due to the specific exposure risk. For example, if prevalence is 1% and 10 individuals are selected at random for quarantine, then on average 0.1 people would be infected. Alternatively, if an index case is isolated upon symptom onset, there would be on average 1.21 individuals infected (for an *R*_0_ = 2.5) prior to symptom onset and potentially identified through contact tracing. With a significant chance of traced contacts being infected, reducing PQT becomes increasingly important. However, traced contacts are likely to enter quarantine earlier in disease (**Fig. S31**). Such an earlier entry necessitates a consequently longer quarantine (generally). Earlier entry makes it more likely that testing early in quarantine will occur during the latent period, when diagnostic sensitivity of the RT-PCR test is highly limited.

Our study is informative for businesses, military operations, and universities, providing quantitative estimation of the residual risk of PQT. The calculated infection risks were used to inform the quarantine and RT-PCR testing strategy deployed by an oil and gas company prior to workers travelling offshore. Of the positive tests obtained under this strategy, 34% were obtained on an exit test following a negative entry test. The exit test prevented 16 infected crew members from exiting quarantine and entering confined quarters offshore while potentially infectious. The results of the time of testing for a given quarantine duration are also useful for public-health decision making when quarantine is required for international, interstate, and social travel.

Our examination of the effects of durations of quarantine and timings of testing is critical to future efforts to balance the risk of PQT with the economic costs, negative impact on mental health, and restrictions on social liberty associated with prolonged quarantines. Timely testing enables a shorter quarantine with equivalent benefits to the much longer 14-day quarantine in prevention of post-quarantine transmission. Our study indicates that the strategy of testing upon entry into quarantine—currently implemented by many institutions and administrative bodies— conveys the least benefit, if infection time is unknown. Testing at exit can provide substantially higher dividends in reducing PQT; or at an optimal timing near 1 week for quarantines of a week or longer. Our result was substantiated both by our integrative analysis of infectivity and diagnostic sensitivity, and by test results demonstrating the utility of tests 96 h into the quarantine of crew members of an offshore oil facility. In determining policies for the duration of travel quarantine and quarantine of traced contacts, full consideration of how timely diagnostic testing aids prevention of post-quarantine transmission is essential to effective and transparent balancing of lives and livelihoods in times of a global pandemic.

## Methods

### Data of SARS CoV-2 tests during quarantine

Between April 11, 2020 and August 26, 2020, there were 4,040 SARS CoV-2 RT-PCR tests conducted among employees of an oil and gas company coming from two regions (stratified by lab location). A third region that was monitored is not included in our data set, as there was low population prevalence entering quarantine and there were no positive tests. During the early stages of the epidemic, both regions used a three-day quarantine with testing on entry. On August 13, employees from region A quarantined at home for seven days, with testing occurring on both entry and exit. While employees were at home, they were asked to practice social distancing in public. Starting on June 25, employees from region B were quarantined in a hotel for five days prior to their departure offshore and tested on both entry and exit. The requirements of an employee to go off-shore were (1) passing the components of a screening form used to filter out symptomatic cases and those potentially exposed, (2) temperature screenings, and (3) completion of the quarantine with no positive RT-PCR test. Upon a positive test, the employee initiated a 14-day isolation period and followed through the company’s case management process. After the isolation period, individuals were able to return back to work contingent upon two negative RT-PCR tests. The use of this data was approved by the Human Participants Review Sub-Committee, York University’s Ethics Review Board (Certificate Number: 2020-323).

### Epidemiological parameters

The average incubation period is 8.29 days ^18^. The latent period (i.e. infected but low probability of infecting contacts) is 2.9 days ^22^. We consider latent periods of 1.9 days and 3.9 days in a scenario analysis ^22^ (**Fig. S11–S19**).

For our baseline analysis, we considered a delay of one day between sample collection and result of RT-PCT test. Thus, the sample is taken one day before the end of quarantine when testing on exit. We also conducted the analysis when there was no delay in testing results to examine the impact on the probability of PQT (**Fig. S20–S23**).

In the baseline analysis, we assumed *R*_0_ = 2.5 and 30.8% of infections are asymptomatic ^8,20^. We further analyzed the scenario in which 22.6% of infections are asymptomatic (**Fig S24–S27**) ^28^. Both of these proportions are consistent with estimates from a systematic meta-analysis ^21^. Asymptomatic infections were assumed to be equally as infectious as symptomatic infections. This assumption is based on measurements of viral loads in asymptomatic infections being comparable to those observed in symptomatic cases ^29,30^.

### Infectivity profile

The infectivity profile has been determined to increase rapidly prior to symptom onset, peak near onset of symptoms,and decrease subsequently ^31^. We specified our infectivity profiles based on the full dataset and R code provided by He et al ^17^, specifying the latent period. The infectivity during the latent period was expressed as exponentially lower (**Supplementary Information: Methods, Infectivity function)**. Imposing the strict threshold where 20 days after symptom onset infectivity is zero ^32–34^ made no significant difference to our estimate of PQT for quarantines of up to 14 days.

### Temporal diagnostic sensitivity of a SARS CoV-2 RT-PCR assay

We utilized the post-symptom onset temporal diagnostic sensitivity for RT-PCR tests of infected individuals ^19^, fitting a logistic regression function to the diagnostic sensitivity data from zero to 25 days post-symptom onset through minimization of least squares. To infer the diagnostic sensitivity prior to symptom onset, we first used this function to perform a slight extrapolation of the diagnostic sensitivity back to the peak, which occurred slightly prior to symptom onset. Second, to determine the diagnostic sensitivity for the remaining portion of the incubation period, we specified the interpolation function determined by the infectivity and the diagnostic sensitivity from post-symptom onset, and used that interpolation function on the pre-symptom onset infectivity to determine pre-symptom onset diagnostic sensitivity (**Supplementary Information: Methods, Diagnostic sensitivity function)**. This process provides the diagnostic sensitivity over the entire course of infection (**Fig. S28**)^13^. We assumed that the specificity of the RT-PCR assay was 100% ^35^.

### Probability of post-quarantine transmission

To calculate the probability of PQT—defined to be the probability of at least one post-quarantine infection—we assumed that the expected post-quarantine transmission is described by a negative binomial distribution with a dispersion parameter of 0.25 ^36^. This value for the dispersion parameter is consistent with numerous published estimates ^37–39^. For sensitivity analyses, we also computed the probability of PQT given Poisson-distributed post-quarantine transmission (**Fig. S29–S30**). In our additional analysis accounting for the underlying prevalence within the community, the probability of PQT was defined as the likelihood that at least one infected individual in a cohort became a source of PQT. Similarly, to calculate the probability of PQT given a negative test on entry for *N* infected individuals, we estimated the probability that at least one of the cases contributed to PQT.

## Data Availability

The number of positive tests and tests conducted at the two regions quarantining the crew members heading offshore are presented in Fig. 2, with other data used in the analysis referenced in Table S1 and in the Methods.

## Data availability

The number of positive tests and tests conducted at the two regions quarantining the crew members heading offshore are presented in **Fig. 2**, with other data used in the analysis referenced in **Table S1** and in the Methods.

## Code availability

The computational code for the analysis was implemented in MATLAB, and it is available at github.com/WellsRC/Optimizing-COVID19-Quarantine-and-Testing-Strategies.

## Author contributions

JPT conceived and designed the study with contributions from other authors, developed the theory and provided initial analyses. CRW derived additional theory, wrote computational code and ran simulations. All authors contributed to interpretation of results, revision of the manuscript and approved the final version of the manuscript.

## Acknowledgements

We thank Justin Abshire for expert data collection. J.P.T. gratefully acknowledges funding from the National Science Foundation grant CCF 1918656, the Elihu endowment, Notsew Orm Sands Foundation, and BHP. G.K., B.S., and R.H.M acknowledge funding from BHP. S.M.M. acknowledges support from the Canadian Institutes of Health Research (grant OV4-170643; Canadian 2019 Novel Coronavirus Rapid Research), the Natural Sciences and Engineering Research Council of Canada, and BHP. A.P.G. gratefully acknowledges funding from NIH UO1-GM087719, the Burnett and Stender families’ endowment, the Notsew Orm Sands Foundation, and BHP.

## Supplementary Information

### Theory

#### Transmission over Time

Transmission of a pathogen from an infected individual is typically time-dependent, based on pathogen shedding and behavioral changes, and can be represented over time by a function *r*(*t*), for which time *t* = 0 represents initial infection. To represent infectiousness, a function *r*(*t*) can be scaled such that

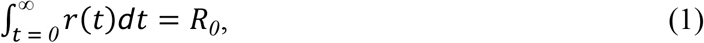

where *R*_*0*_is the basic reproduction number: the expected number of infections consequent to a single infected individual under a scenario of no intervention. Specifying a discrete end to the infection at time *t*_*e*_ such that *r*(*t*) = *0* for *t* > *t*_*e*_,

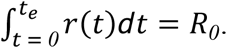

Infectiousness during discrete timespans *t*_*2*_ − *t*_*1*_ (e.g. days) can be calculated as

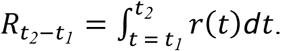

#### Self-isolation at Symptom Onset

A significant means of intervention to prevent infection is self-isolation of infected individuals upon symptom onset. The expected effect on onward transmission of an intervention such as self-isolation of a case that becomes symptomatic at time *t*_*s*_ can be calculated as

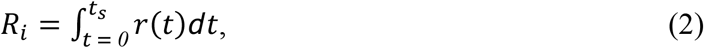

provided that all individuals self-isolate upon presentation with symptoms. If 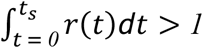, then even perfect self-isolation upon symptom onset will be insufficient to extinguish disease transmission. We express the transmission over time for a symptomatic individual who isolates upon symptom onset as

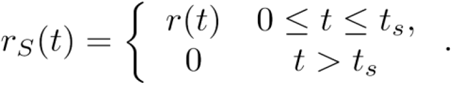

If the outcome of infections leads to a proportion of infected individuals *p*_*a*_that can infect others but that never manifest symptoms (i.e. that are asymptomatic carriers), then transmission may be partitioned into the contributions of symptomatic and asymptomatic cases as *R*_*0*_ = *R*_*0,s*_*p*_*s*_ + *R*_*0,a*_*p*_*a*_, in which the probability of a symptomatic case *p*_*s*_ = (*1* − *p*_*a*_). *R*_*0,s*_ and *R*_*0,a*_ can be equated to distinct infectiousness functions *r*_*s*_(*t*) and *r*_*a*_(*t*), in the absence of self-isolation. For simplicity of presentation in ensuing theory, it will be assumed that *R*_*0,s*_ = *R*_*0,a*_ and the same infectivity profile in the absence of self-isolation (i.e. *r*_*s*_(*t*) = *r*_*s*_(*t*) = *r*(*t*)) ^1,2^. Alternate overall transmission and alternate forms of infectivity over time for asymptomatic cases may easily be partitioned and tracked in the theory that follows should there be evidence to substantiate their difference.

The presence of asymptomatic carriers increases the degree of transmission consequent to a self-isolation intervention from that shown by **Eq. 2** to

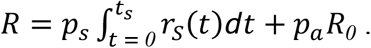

#### Quarantine

##### Quarantine with a Known Time of Infection

A longstanding approach to limit disease spread is the quarantine of individuals *who have no prior indication of potential for disease* but intend to migrate from a population in which there is current transmission to a population with lower or zero disease prevalence. Because quarantined individuals experience a significant restriction of personal freedom, it is important to minimize the duration of quarantine *q*, but also maximize its effectiveness in limiting post-quarantine transmission. Quarantine of *q* days from time *t*_*q*_to time *t*_*q*_ + *q* limits total expected post-quarantine transmission to

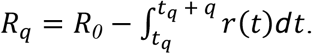

For policy decision-making regarding quarantine duration, the expected post-quarantine transmission is typically most important, and can be calculated as

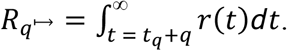

If individuals self-isolate, there is a trivial case in which *t*_*s*_ ≤ *t*_*q*_ + *q* and 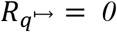; otherwise, *t*_*s*_ > *t*_*q*_ + *q* and

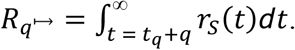

Including asymptomatic carriers,

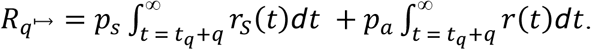

Unfortunately, these expressions are unlikely to be useful in this form for quantifying the benefits of quarantine in reducing transmission. In the case of quarantine of migrants from one population to another, the time of infection—and correspondingly the time of quarantine *t*_*q*_—are rarely known.

##### Quarantine with an Unknown Time of Infection

In a rapidly spreading epidemic, individuals who might be entering quarantine will tend to be early in disease time course. In a rapidly declining epidemic, individuals who might be entering quarantine will tend to be later in disease time course. In a steady-state epidemic with case counts 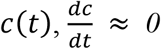 over the period from *t*_0_ to *t*_s_ such that individuals entering quarantine are evenly distributed across the disease time course. Provided all individuals experience symptoms at time *t*_*s*_ that qualify them for isolation instead of quarantine, then the expected post-quarantine infectivity is

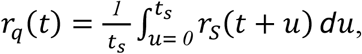

and expected post-quarantine transmission from an infected individual is

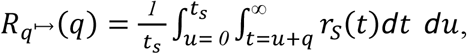

a function of days of quarantine *q*. For asymptomatic carriers entering within disease time course *t*_*e*_,

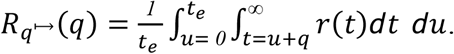

Incorporating both symptomatic and asymptomatic infections,

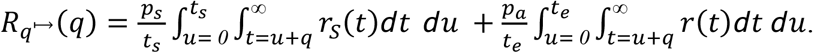

A similar approach that incorporates symptomatic and asymptomatic cases by their proportions within the population may be performed throughout the rest of the scenarios below, and will not be specifically pointed out for each scenario.

#### Testing

##### Testing with a Known Time of Infection

Diagnostic test sensitivity *s*(*t*) is also time-dependent. Assaying for components of the pathogen (e.g. DNA, RNA, or protein), diagnostic sensitivity typically is zero to low very early in disease before the pathogen load burgeons, then declines in the later stages of disease when immune responses develop and infection is suppressed (**Fig. S28**). In a disease for which tests can diagnose infections during the incubation phase, testing can enhance the efficacy of quarantine by identifying individuals to be isolated instead of quarantined, thereby preventing future transmission from cases that persist as infectious through an earlier exit from quarantine than would be called for in case isolation.

##### Testing with an Unknown Time of Infection

The temporal diagnostic sensitivity of a test for infected cases with an unknown time of infection can be calculated by integrating over the unknown time of infection, such that

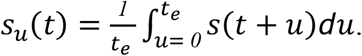

#### Quarantine and Testing

##### Quarantine with an Unknown Time of Infection with Testing on Entry

Assuming the duration of the quarantine, *q*, is longer than the delay between administering the test and acting to isolate upon a positive result, the expected post-quarantine infectivity over time of a symptomatic individual whose time of infection is unknown and who is tested for disease on entry to quarantine is

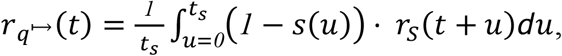

in terms of time from infection. In terms of *q* days of quarantine, the expected post-quarantine transmission is

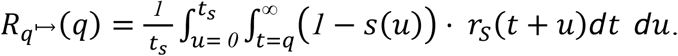

For asymptomatic carriers,

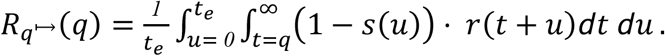

##### Quarantine with an Unknown Time of Infection with Testing on Entry and Exit

Expected post-quarantine transmission from an individual whose time of infection is unknown and who is tested for disease upon entry and at the last opportunity prior to the end of quarantine is

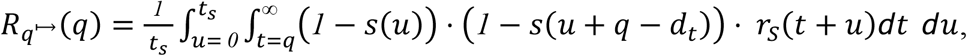

where *d*_*t*_ is the delay between administering the test and isolation if positive. For asymptomatic carriers,

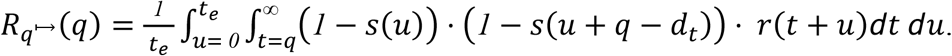

##### Quarantine with Testing at Any Time(s)

Expected post-quarantine transmission of an infected individual whose time of infection is unknown and who is tested for disease at any time *0* ≤ *t*_*t*_ ≤ *q* − *d*_*t*_ is

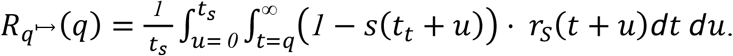

For asymptomatic carriers,

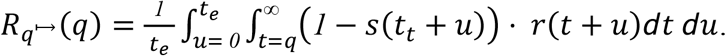

Additional terms (*1* − *s*(*u* + *t*_*k*_)), where *k* indexes testing times, may be included as terms within the product inside the double integral to quantify the expected post-quarantine transmission of any schedule of testing to be applied during quarantine.

##### Quarantine with a Negative Test on Entry

The probability density for obtaining a false negative upon entry for a symptomatic individual is

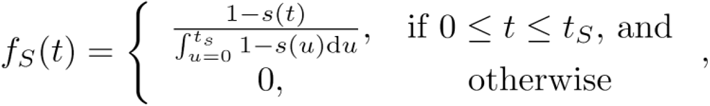

and the probability density for an asymptomatic individual is

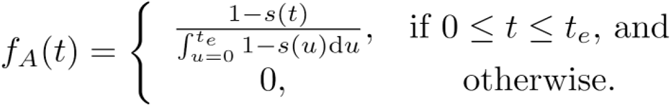

The expected post-quarantine infectivity over time of a symptomatic individual who tested negative for disease on entry to quarantine is

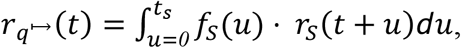

in terms of time from infection. In terms of *q* days of quarantine, the expected post-quarantine transmission is

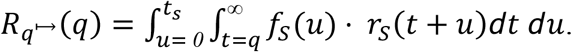

For asymptomatic carriers, the expected post-quarantine infectivity is

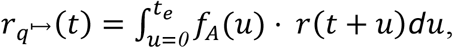

and the expected post-quarantine transmission is

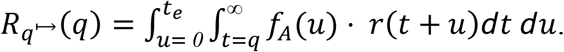

#### Contact Tracing

Tracing of individuals who have had contact with an index case identifies persons whose quarantine would reduce the risk of disease transmission from recently exposed individuals. When an individual is identified as a contact of an index case, the expected time of infection is not the same as that of an individual selected at random from an infected population. Restricting our attention to transmissions occurring between an index case and their contacts, there are four nominal transmission relationships to be considered, of which three are considered relevant to an attentive program of contact tracing and quarantine (**Table S2**): the asymptomatic or pre-symptomatic contact may have infected the index case, or may have been infected by the index case. Here we excluded from calculation the case in which a pre-symptomatic individual infects the index case, because that scenario is formally impossible with a fixed *t*_*s*_ and rigorous self-isolation and self-identification upon symptoms, and unlikely even with variable *t*_*s*_ and imperfect adherence to self-isolation and self-identification.

**Table S2.** Modelled infectivity functions for the contact during tracing.

| Contact | Infected by the index case | Infected the index case |
| --- | --- | --- |
| Symptomatic case | $r_{I \rightarrow S}(t)$ | — |
| Asymptomatic carrier | $r_{I \rightarrow A}(t)$ | $r_{A \rightarrow I}(t)$ |

##### A To-be-Symptomatic Contact Infected by the Index Case but not yet Symptomatic

By assumption, infection of the contact must have occurred prior to the onset of symptoms in the index case. The likelihood that an infection from the index case occurred at a time during the disease time course of the index case should proportionally follow *r*(*t*) (**Eq. 1**). Thus, the probability density for infection—on the timescale *t* of the infection of the index case that was identified at symptom onset—is

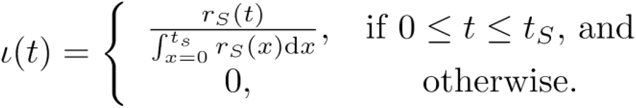

The probability density for the time since infection of the to-be-symptomatic contact—on the timescale *t* of the contact—is

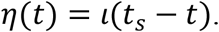

Thus, the erstwhile expected infectivity from the contact that was infected by the index case from the time of intervention by a quarantine is

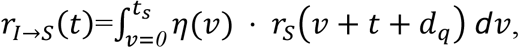

where *d*_*q*_ is the delay from identifying the index case to quarantine of the contact. The expected post-quarantine transmission by the contact after a quarantine of duration *q* is

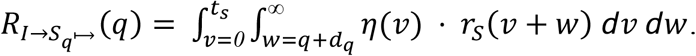

##### An Asymptomatic Carrier Contact Infected by the Index Case

The expected infectivity of an asymptomatic contact infected by the index case—from time *t* = 0 at intervention by quarantine—is

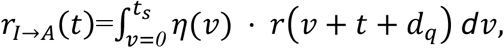

where *d*_*q*_ is the delay from identifying the index case to quarantine of the contact. The expected post-quarantine transmission from the asymptomatic contact infected by the index case starting from the time of intervention by a quarantine of duration *q* is

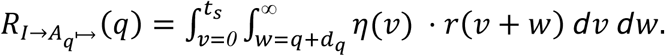

##### An Asymptomatic Contact that Infected the Index Case

Because the index case was assumed to be identified due to symptom onset, an asymptomatic contact that infected the index case must have already been infected for a duration of at least *t*_*s*_ + *d*_*q*_. Consequently, the probability density of infection from that contact is

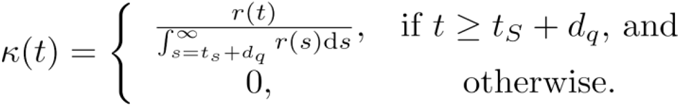

Setting 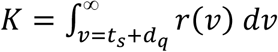, the expected infectivity of the asymptomatic contact that infected the symptomatic index case—from time *t* = 0 at intervention by quarantine—is

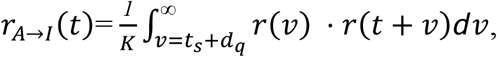

and the expected post-quarantine transmission is

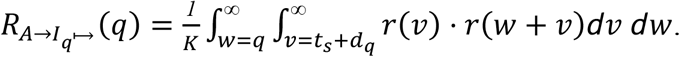

Continuing our assumption that individuals are assiduously self-isolating upon symptom onset and recalling that 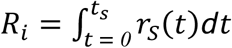 (**Eq. 2**), we can tabulate the expected transmission by contacts of the index that are classified into three kinds (**Table S3**). By assumption, a contact to become symptomatic could not have infected the index case, because otherwise in an assiduously self-isolating population, that contact would have been the index case.

**Table S3.** Expected infections from contacts of each modeled transmission type.

| Contact | Infected by the index case | Infected the index case |
| --- | --- | --- |
| Symptomatic case | $p_s R_i$ | — |
| Asymptomatic carrier | $p_a R_i$ | $p_a R_o$ |

Combining all three transmission functions of contacts of an index case discovered due to appearance of symptoms, the expected post-quarantine infectivity

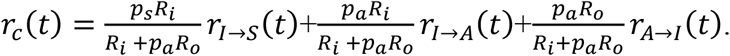

Incorporating a quarantine of duration *q* for contacts, the expected post-quarantine transmission

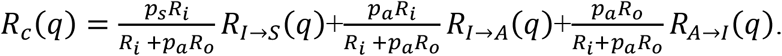

#### Probability of post-quarantine transmission

The probability of post-quarantine transmission is specified to be the probability that an infected individual exits quarantine, but can still infect one or more individuals. We calculated this probability under a negative-binomial model appropriate when superspreaders play a role in transmission, as well as a Poisson distribution appropriate when transmission is fairly evenly distributed among infected individuals.

##### Negative-binomial distribution

We specified a negative binomial distribution

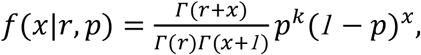

with dispersion parameter *k* = 0.25 ^3^ and 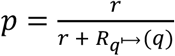 such that the average of the distribution was 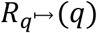. Thus, the corresponding probability of post-quarantine transmission with negative binomially-distributed transmissions from a case is

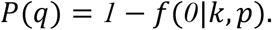

##### Poisson distribution

Specifying a Poisson distribution producing an expected number of secondary infections post-quarantine transmission of 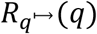, the probability of transmission after a quarantine of duration *q* days

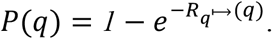

### Population prevalence

Given a cohort size *N* and a prevalence of *ρ*, the probability of post-quarantine transmission is *1* − (*1* − *P*(*q*))^*Nρ*^.

### Methods

#### Infectivity function

We use a Gamma function to specify the infectivity over the disease time course (**Fig. S1 and Fig. S11**). We generated the infectivity profile during the pre-symptomatic phase for each duration of the pre-symptomatic period corresponding to each latent period, using the R code provided from He et al ^4^. However, as a matter of accounting for the full disease time course, a level of infectivity during the latent period prior to the discrete onset of the distribution provided by He et al must also be specified. Therefore, we specified the infectivity during this early period of infection as *A*(*e*^*m*·(*t*)^ − *1*), where the constants *m* and *A* are estimated such that the infectivity function *r*(*t*) is smooth and continuous over the entire disease time course. Since the infectivity profile after the latent stage is described by a Gamma function (which has an initial value of zero), we truncate the exponential function at time *t*_*L*_ + Δ*t*, where *t*_*L*_ is the duration of the latent period; Δ*t* was set as the difference between *t*_*L*_ and the upper bound of *t*_*L*_ (where the difference in the log-likelihood at *t*_*L*_ and at *t*_*L*_ + Δ*t* was 1.96 ^5^).

#### Diagnostic sensitivity function

To characterize the diagnostic sensitivity post-symptom onset, we estimated the coefficients of a logistic regression model

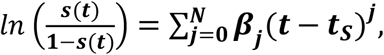

by fitting the function *s*(*t*) to diagnostic test-sensitivity data from day zero to 25 days post-symptom onset ^6^ through the minimization of least squares

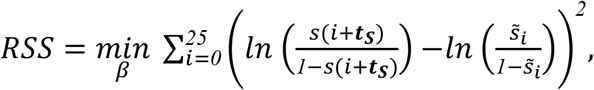

where 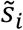 denotes the observed diagnostic sensitivity at day *i* post-symptom onset. The peak infectivity occurs prior to symptom onset from the inferred infectivity curves ^4^, implying that the infectivity curve is monotonically decreasing over time after symptom onset. To be consistent, the sensitivity should also be monotonically decreasing over time after symptom onset as infectivity (a proxy for the viral load) is decreasing. Therefore, a constraint that the maximum sensitivity after symptom onset occurred at time zero was included in the estimation of the coefficients of the logistic regression model.

To select the number of coefficients in the logistic regression model, we used the Akaike information criterion,

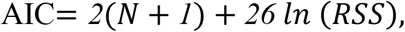

where there are *N* + 1 coefficients being estimated for the 26 data points. The logistic regression model with the lowest AIC value was used to determine the diagnostic sensitivity.

We used diagnostic test-sensitivity data from zero to 25 days post-symptom onset ^6^ and the infectivity profile post-symptom onset ^4^ to construct a mapping from infectivity to diagnostic sensitivity, then used that mapping to infer the diagnostic sensitivity during the incubation period from the infectivity pre-symptom onset. To infer the diagnostic sensitivity during the unobserved incubation period, we defined an interpolation function for the diagnostic sensitivity based on the Cartesian pairing of *r*(*t*) and *s*(*t*) from symptom onset. Since the peak of infectivity occurred prior to symptom onset, we performed a slight extrapolation of the function *s*(*t*) determined by logistic regression. This extrapolation lies within a small range between the symptom-onset diagnostic sensitivity of 0.96 and an upper limit of 1.0 for each latent period considered, so that our results are not sensitive to this extrapolation.

## Supplementary Tables

**Table S1:** Parameter descriptions and values used to assess quarantine and testing strategies

| Description | Parameter | Value | Reference |
| --- | --- | --- | --- |
| Basic reproductive number | $R_0$ | 2.5 and 2.0 | 7 |
| Basic reproductive number for symptomatic infection | $R_{0,s}$ | $R_0$ | 2 |
| Basic reproductive number for asymptomatic infection | $R_{0,a}$ | $R_0$ | 2 |
| Incubation period | $t_S$ | 8.29 days | 8 |
| Duration of disease in asymptomatic individuals | $t_e$ | $t_S + 20$ days | 9–11 |
| Proportion of infections that are asymptomatic | $p_a$ | 30.8%<br>22.6% | 12,13<br>13,14 |
| Latent period | $t_L$ | 2.9<br>1.9 and 3.9 | 15 |

## Supplementary Figures

**Figure S1.**
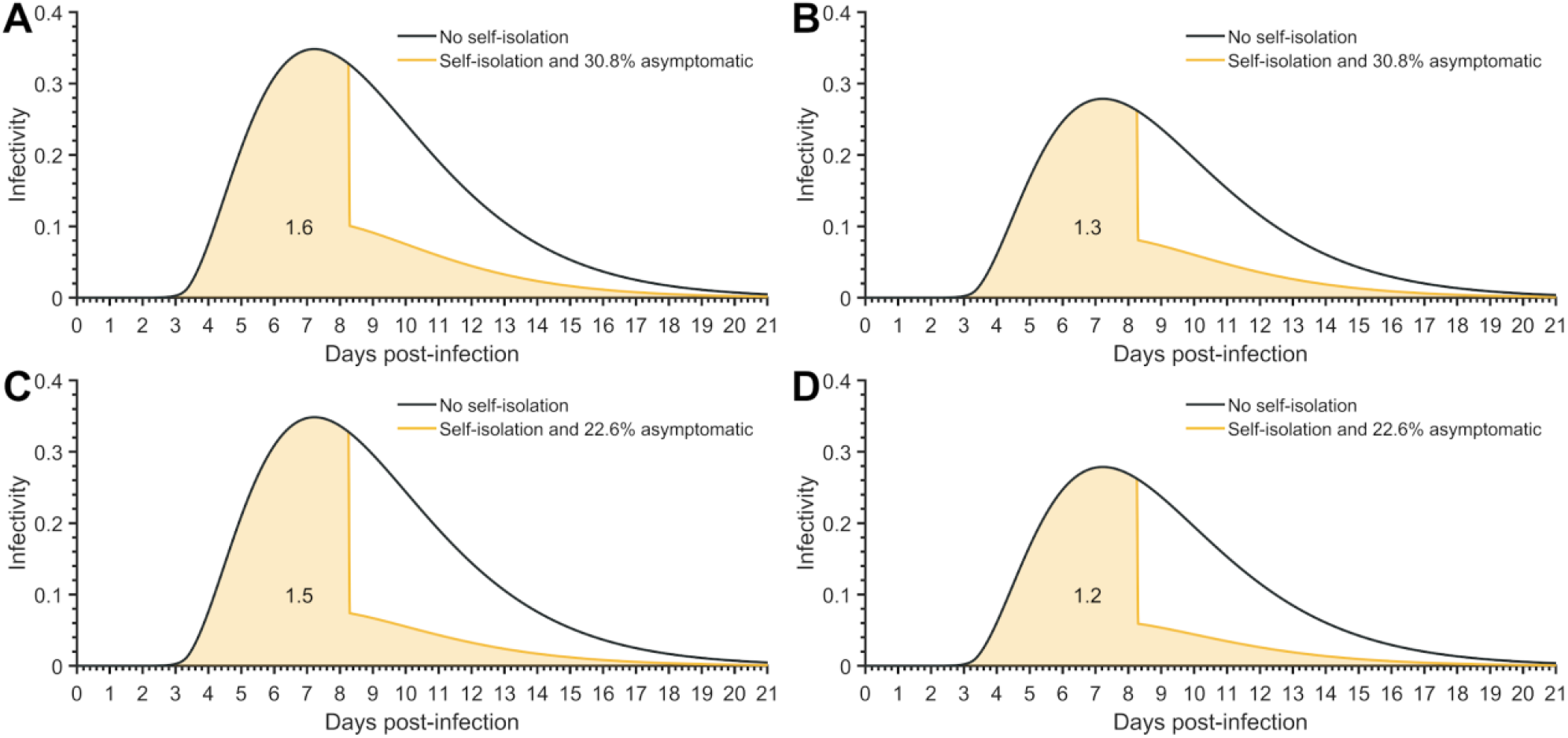
Average infectivity profile for a known time of infection under no self-isolation upon symptom onset (black) and perfect isolation upon symptom onset (yellow line) for (**A**) *R*_0_ = 2.5 and 30.8% of infections being asymptomatic (resulting in 1.6 secondary infections, yellow fill), (**B**) *R*_0_ = 2 and 30.8% of infections being asymptomatic (resulting in 1.3 secondary infections, yellow fill), (**C**) *R*_0_ = 2.5 and 22.6% of infections being asymptomatic (resulting in 1.5 secondary infections, yellow fill) and (**D**) *R*_0_ = 2 and 22.6% of infections being asymptomatic (resulting in 1.2 secondary infections, yellow fill).

**Figure S2:**
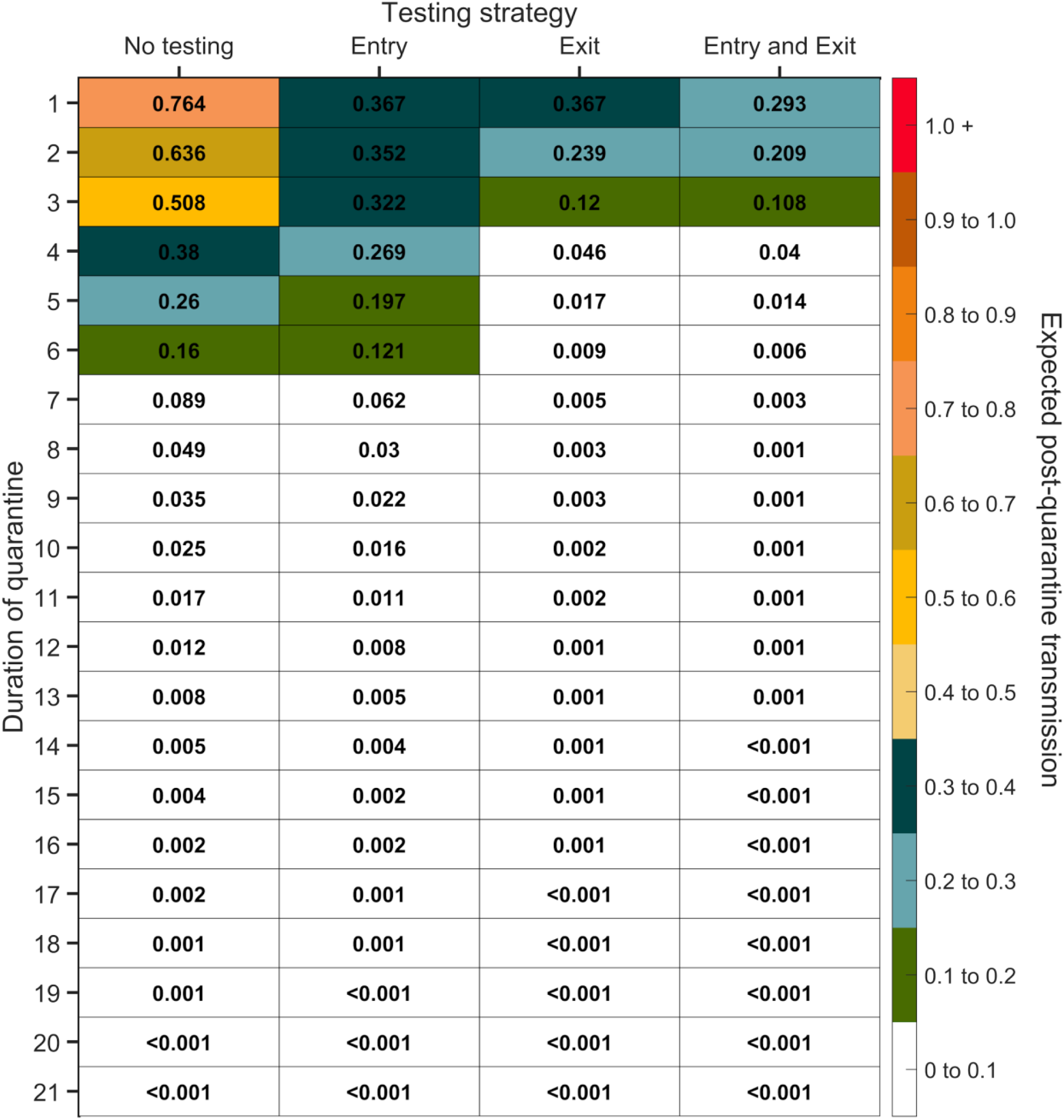
Expected post-quarantine infections for durations of quarantine of 1–21 days, with an incubation period of 8.29 days, a latent period of 2.9 days, 30.8% of infections being asymptomatic, perfect self-isolation of symptomatic infections when symptomatic, uniform entry within the incubation period by symptomatic cases, and uniform entry across the disease time course for asymptomatic cases, with no testing, testing on entry, testing on exit, and testing on entry and exit. Because of the time required to obtain test results, sampling for the test on exit was assumed to occur the day before the quarantine was completed. Cells that share a background color in common indicate equivalent durations of quarantine associated with each of the testing strategies.

**Figure S3:**
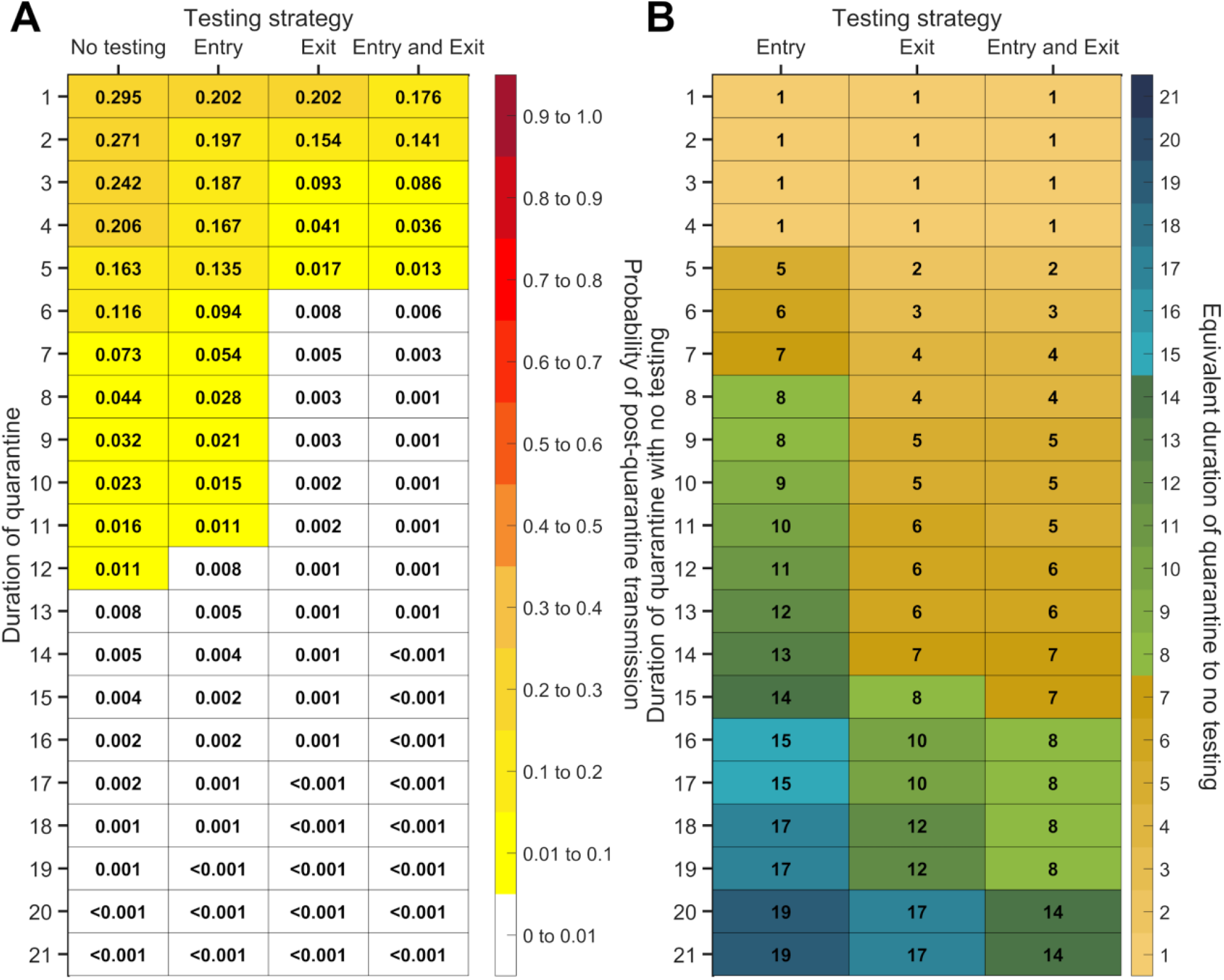
For durations of quarantine from 1–21 days, when a symptomatic individual enters quarantine uniformly within the incubation period and asymptomatic individuals enter uniformly across the disease time course, with an incubation period of 8.29 days, a latent period of 2.9 days, with 30.8% of infections being asymptomatic, and perfect self-isolation of symptomatic infections, (**A**) the probability of post-quarantine transmission (probability of one or more post-quarantine infections) with no testing, when tested upon entry to quarantine, when tested on exit from quarantine, and when tested on entry and exit from quarantine, and (**B**) the durations of quarantine with testing on entry, testing on exit, and testing on entry and exit that perform just as well or better than a quarantine with no testing. Because of the time required to obtain test results, sampling for the test on exit is assumed to occur the day before the quarantine is complete. Cells that share a background color in common indicate equivalent durations of quarantine associated with each of the testing strategies.

**Figure S4:**
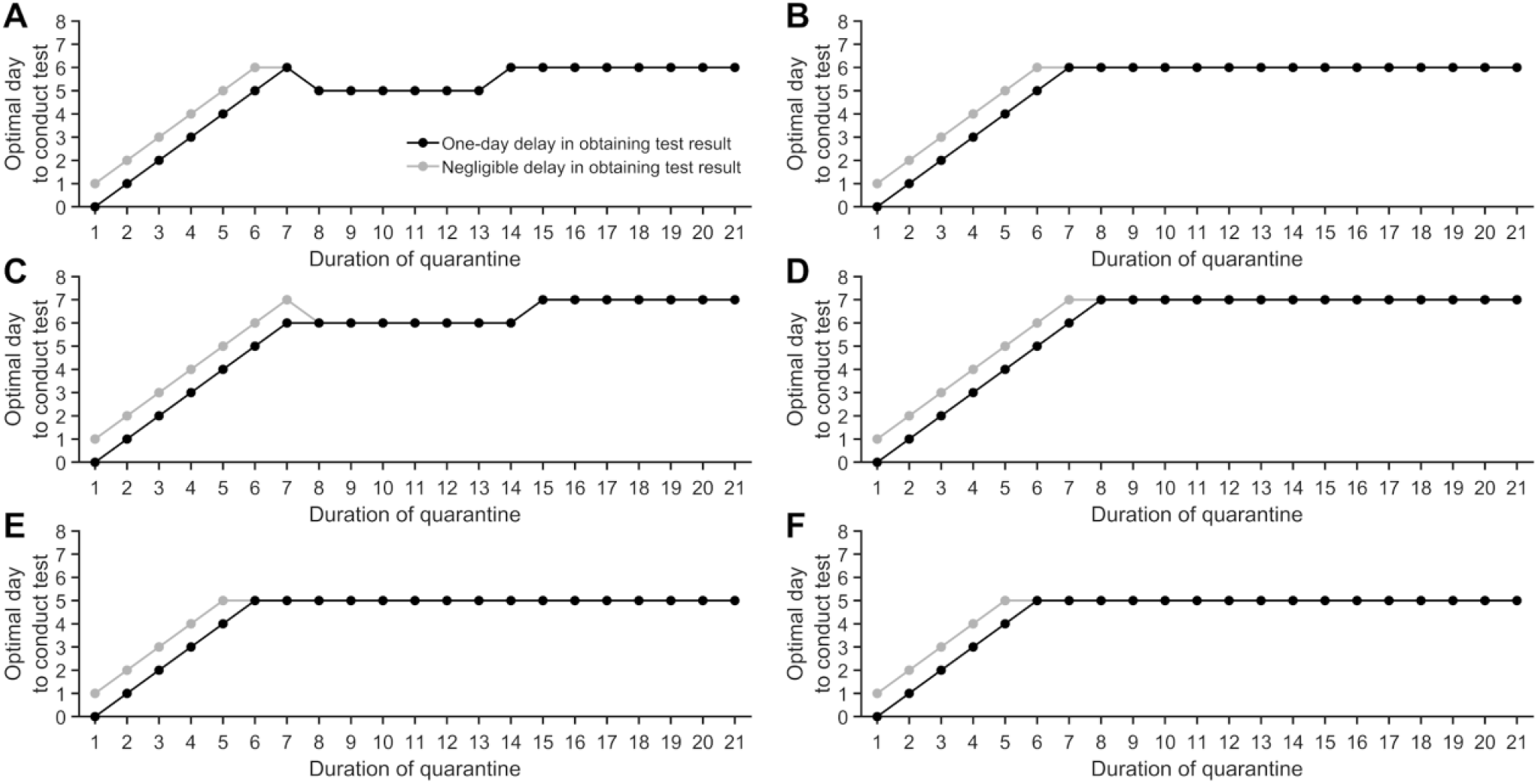
With 30.8% of infections asymptomatic, perfect self-isolation of symptomatic infections, and an incubation period of 8.29 days, the optimal day of testing to obtain the minimum post-quarantine transmission specifying a latent period of (**A**) 2.9 days with uniform entry into quarantine, (**B**) 2.9 days and entry into quarantine as a traced contact, (**C**) 1.9 days and uniform entry into quarantine, (**D**) 1.9 days and entry into quarantine as a traced contact, (**E**) 3.9 days and uniform entry into quarantine, and (**F**) 3.9 days and entry into quarantine as a traced contact for a one-day delay in obtaining test results (black) and a negligible delay in obtaining test results (gray).

**Figure S5:**
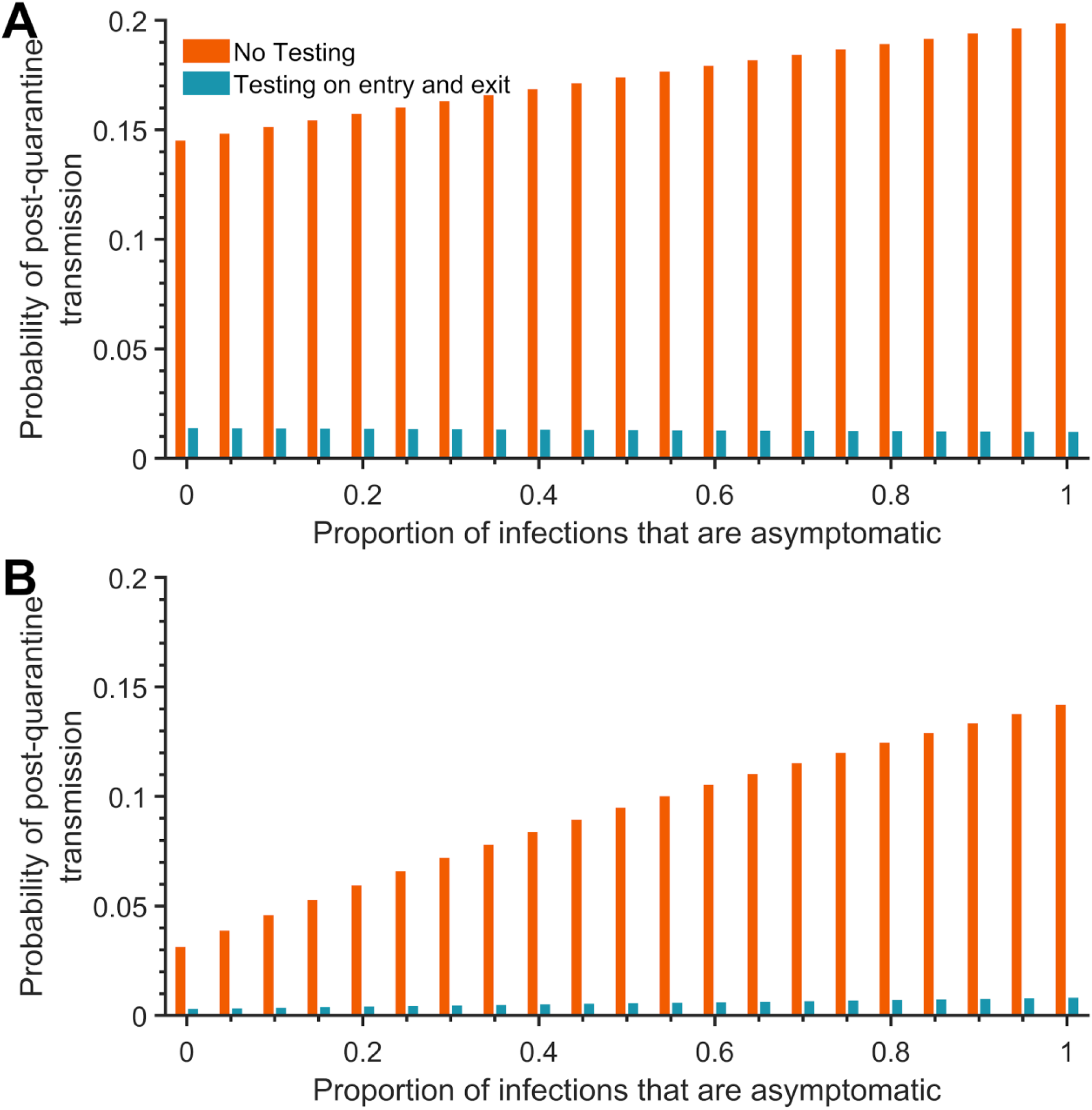
With perfect self-isolation of symptomatic infections, an incubation period of 8.29 days and a latent period of 2.9 days, and proportions of from 0–1 of infections being asymptomatic, the probability of post-quarantine transmission (probability of one or more post-quarantine infections) when symptomatic individuals enter quarantine uniformly within the incubation period and asymptomatic individuals enter uniformly across the disease time course, with no testing (red) and when tested on entry and exit from quarantine (blue) for a (**A**) five-day quarantine and a (**B**) seven-day quarantine. The exit test was assumed to occur 96 h after entry into quarantine.

**Figure S6.**
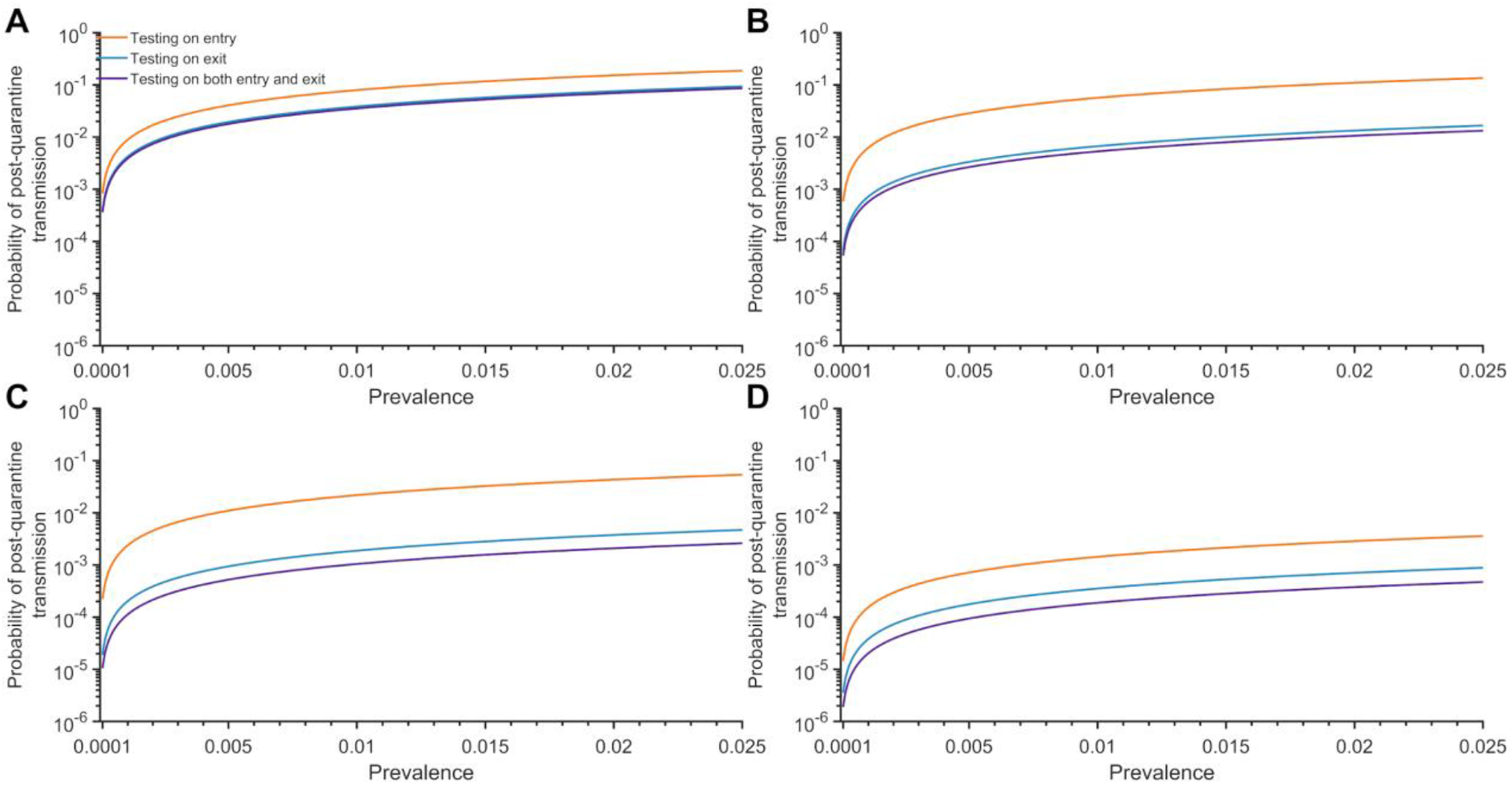
Specifying an incubation period of 8.29 days and a latent period of 2.9 days, the probability of post-quarantine transmission accounting for underlying community prevalence in a cohort (crew) of 40 employees for testing on entry (orange), testing on exit (blue), and testing on both entry and exit (purple) for a (**A**) three-day quarantine, (**B**) five-day quarantine, (**C**) seven-day quarantine, and (**D**) 14-day quarantine. Because of the time required to obtain test results, sampling for the test on exit is assumed to occur the day before the quarantine is complete.

**Figure S7:**
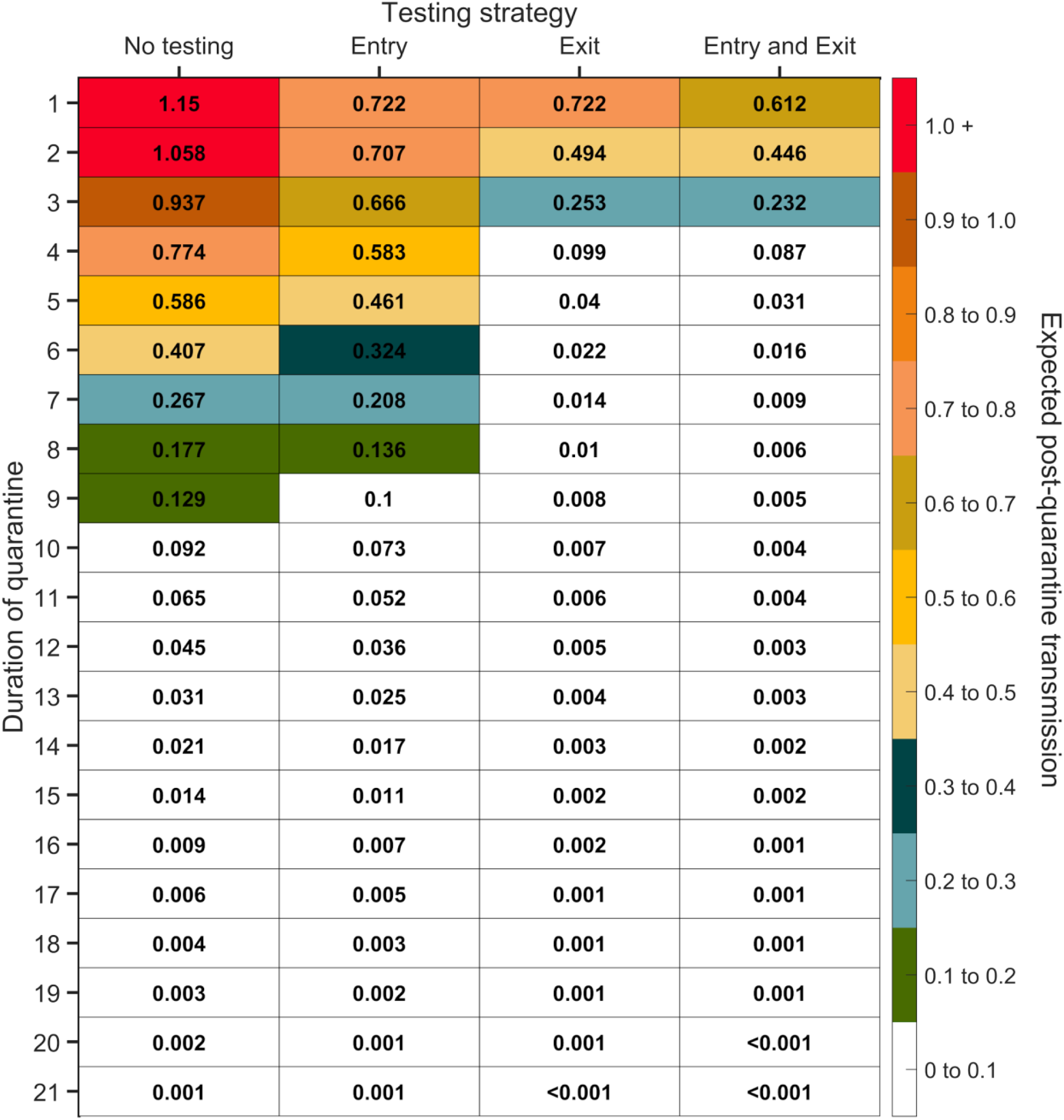
Expected post-quarantine infections for durations of quarantine of 1–21 days, with an incubation period of 8.29 days, a latent period of 2.9 days, 30.8% of infections being asymptomatic, perfect self-isolation of symptomatic infections when symptomatic, and entry through contact tracing, with no testing, testing on entry, testing on exit, and testing on entry and exit. Because of the time required to obtain test results, sampling for the test on exit was assumed to occur the day before the quarantine was completed. Cells that share a background color in common indicate equivalent durations of quarantine associated with each of the testing strategies.

**Figure S8:**
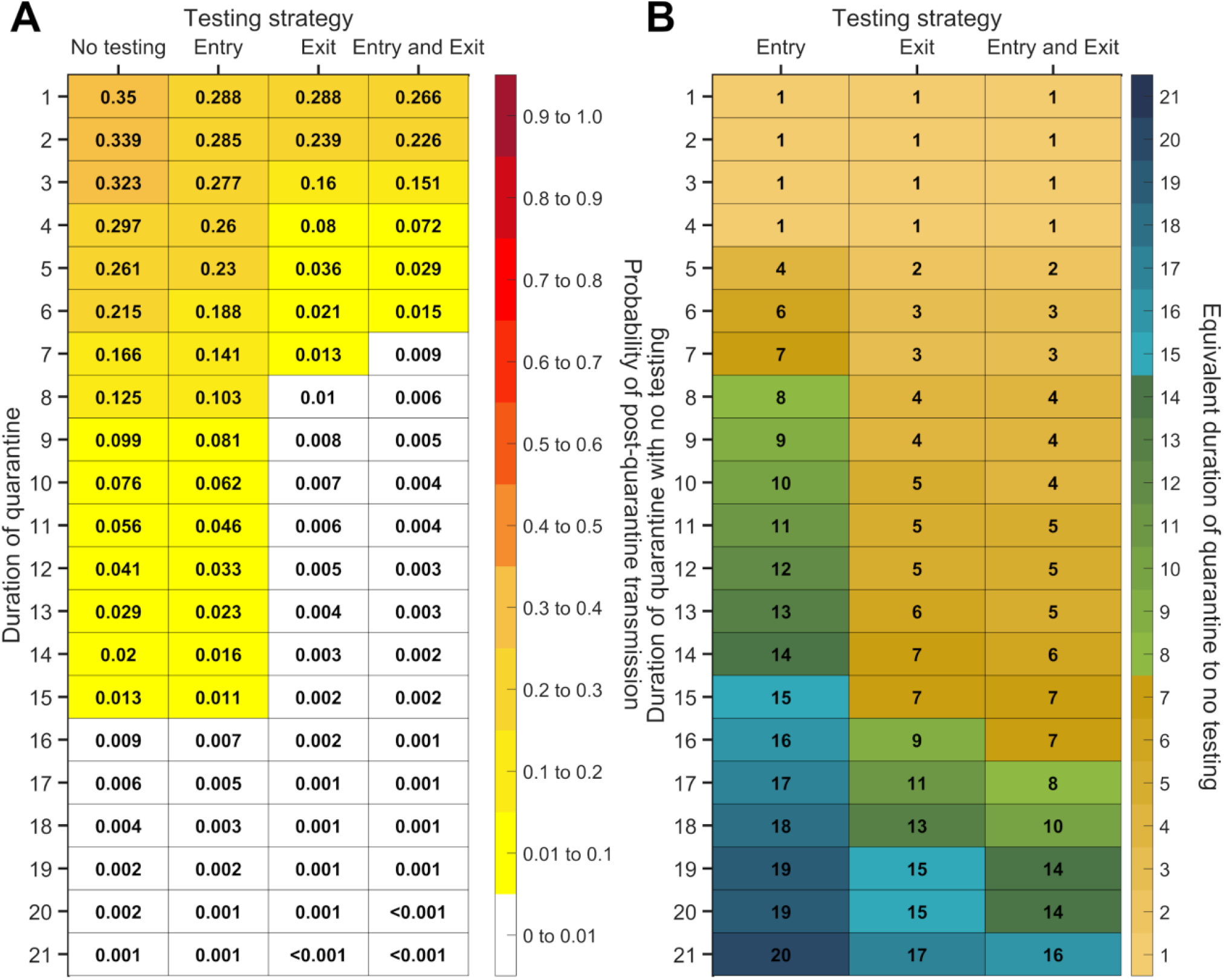
For durations of quarantine from 1–21 days, when an individual enters quarantine through contact tracing, with an incubation period of 8.29 days, a latent period of 2.9 days, with 30.8% of infections being asymptomatic, and perfect self-isolation of symptomatic infections, (**A**) the probability of post-quarantine transmission (probability of one or more post-quarantine infections) with no testing, when tested upon entry to quarantine, when tested on exit from quarantine, and when tested on entry and exit from quarantine, and (**B**) the durations of quarantine with testing on entry, testing on exit, and testing on entry and exit that perform just as well or better than a quarantine with no testing. Because of the time required to obtain test results, sampling for the test on exit is assumed to occur the day before the quarantine is complete. Cells that share a background color in common indicate equivalent durations of quarantine associated with each of the testing strategies.

**Figure S9:**
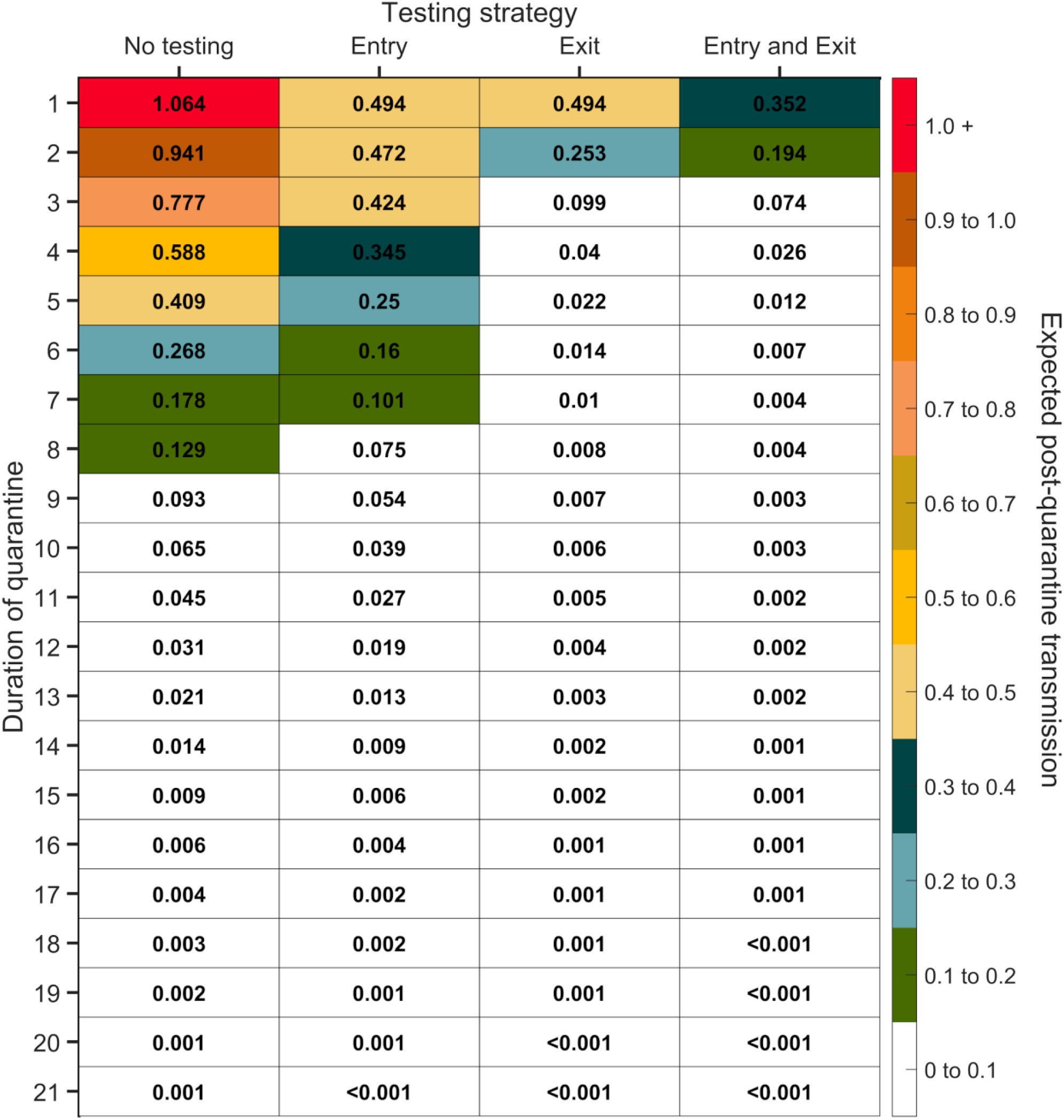
Expected post-quarantine infections for durations of quarantine of 1–21 days, with an incubation period of 8.29 days, a latent period of 2.9 days, 30.8% of infections being asymptomatic, perfect self-isolation of symptomatic infections when symptomatic, and entry through contact tracing with a one-day tracing delay, with no testing, testing on entry, testing on exit, and testing on entry and exit. Because of the time required to obtain test results, sampling for the test on exit was assumed to occur the day before the quarantine was completed. Cells that share a background color in common indicate equivalent durations of quarantine associated with each of the testing strategies.

**Figure S10:**
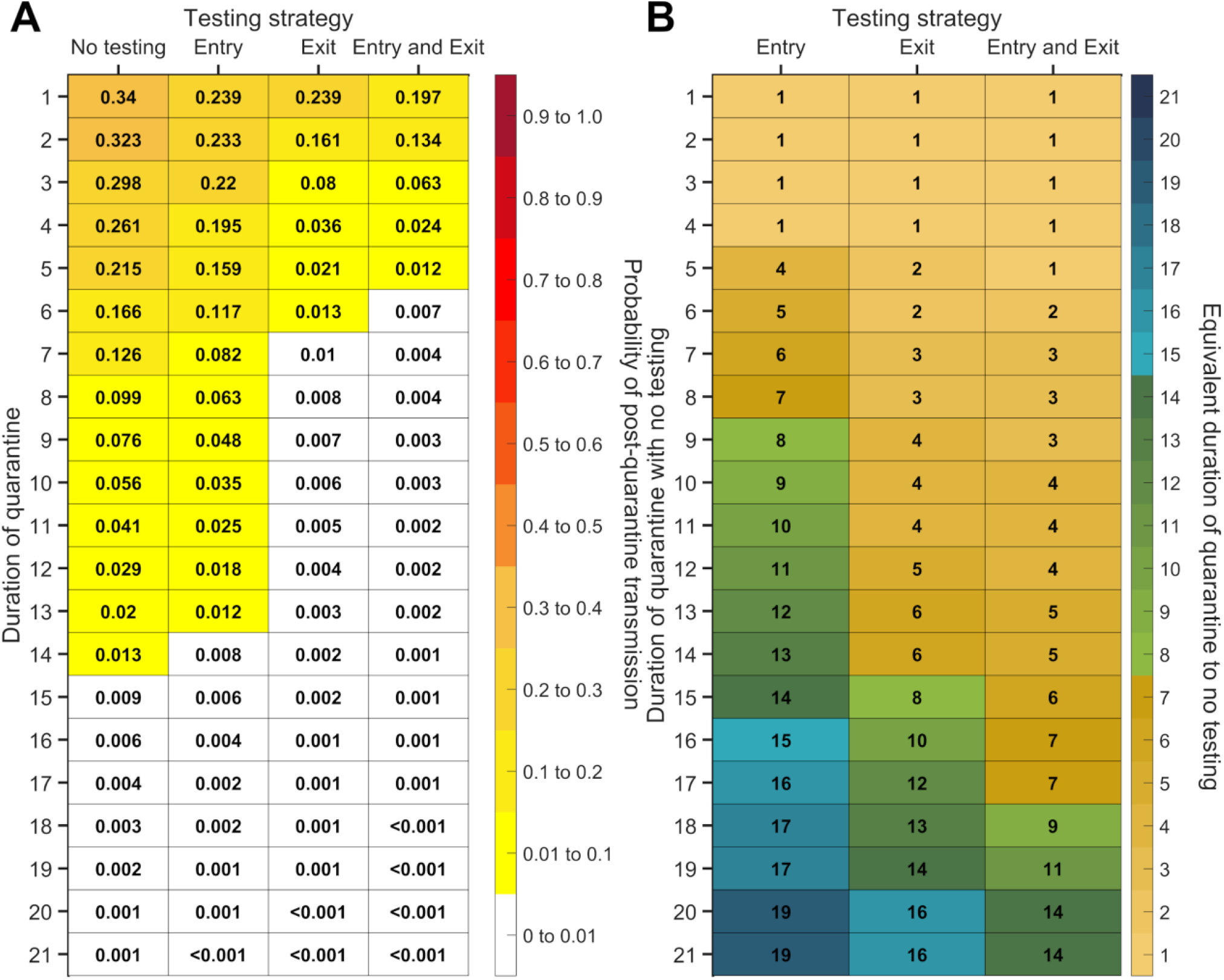
For durations of quarantine from 1–21 days, when an individual enters quarantine through contact tracing with a one-day tracing delay, with an incubation period of 8.29 days, a latent period of 2.9 days, with 30.8% of infections being asymptomatic, and perfect self-isolation of symptomatic infections, (**A**) the probability of post-quarantine transmission (probability of one or more post-quarantine infections) with no testing, when tested upon entry to quarantine, when tested on exit from quarantine, and when tested on entry and exit from quarantine, and (**B**) the durations of quarantine with testing on entry, testing on exit, and testing on entry and exit that perform just as well or better than a quarantine with no testing. Because of the time required to obtain test results, sampling for the test on exit is assumed to occur the day before the quarantine is complete. Cells that share a background color in common indicate equivalent durations of quarantine associated with each of the testing strategies.

**Figure S11:**
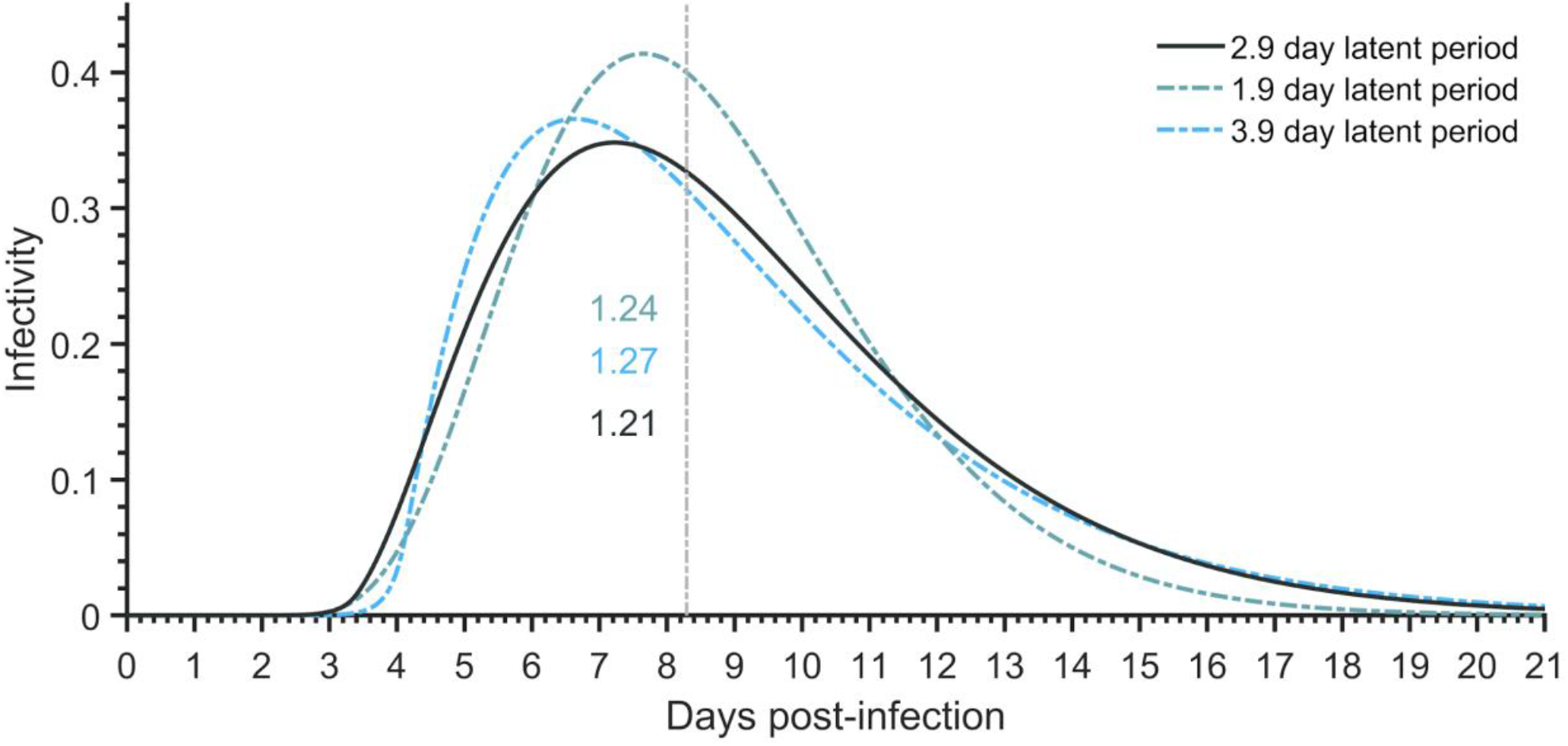
Infectivity profiles of an individual for an incubation period of 8.29 days and assuming no self-isolation upon symptom onset, corresponding to the reported duration of the latent period (2.9, black), and latent periods one day longer (3.9, dashed blue), and one day shorter (1.9, dashed green), and numbers of secondary infections that occur within the incubation period for a 2.9-day latent period (1.21, black), for a 3.9-day latent period (1.27, blue), and for a 1.9-day latent period (1.24, green).

**Figure S12:**
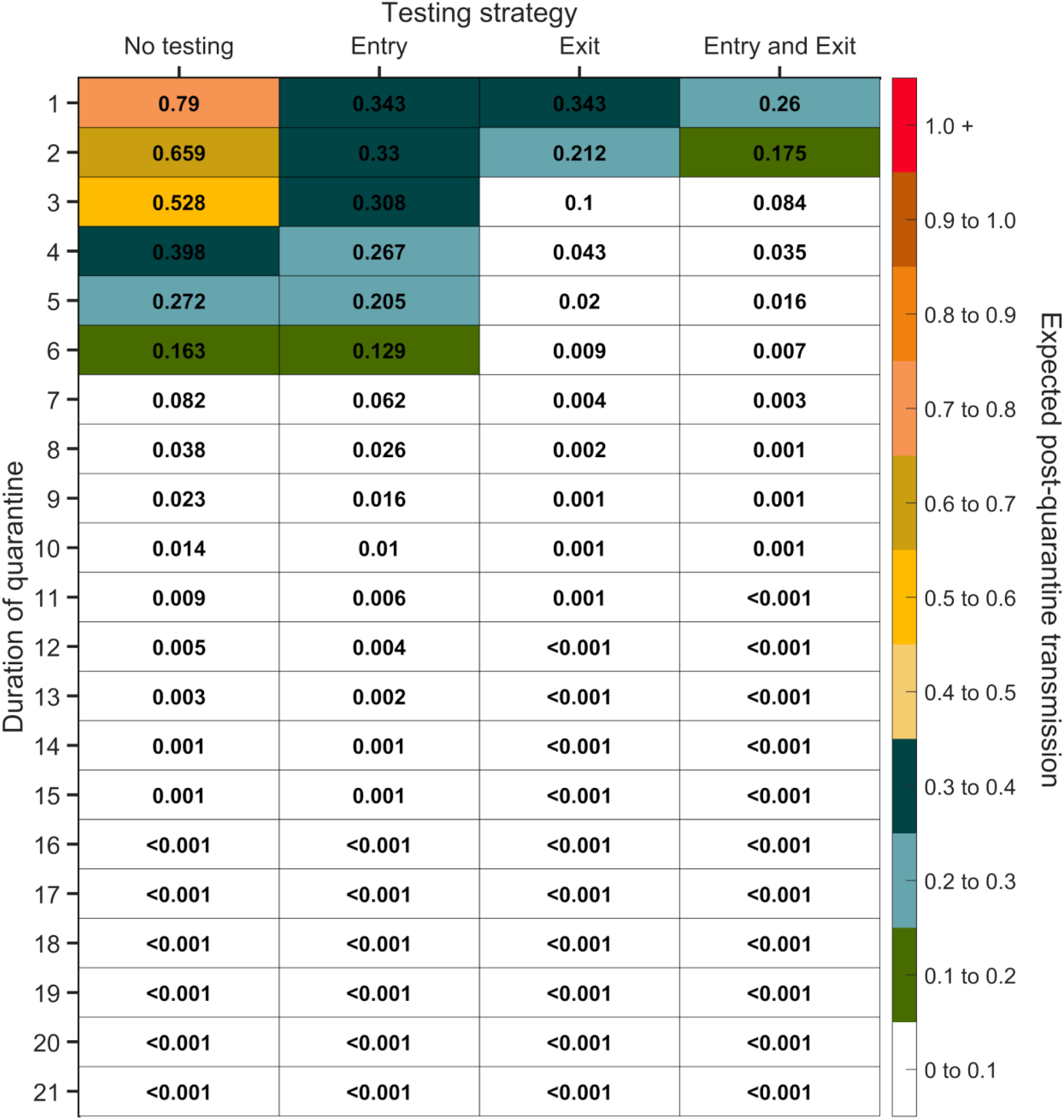
Expected post-quarantine infections for durations of quarantine of 1–21 days, with an incubation period of 8.29 days, a latent period of 1.9 days, 30.8% of infections being asymptomatic, perfect self-isolation of symptomatic infections when symptomatic, uniform entry within the incubation period by symptomatic cases, and uniform entry across the disease time course for asymptomatic cases, with no testing, testing on entry, testing on exit, and testing on entry and exit. Because of the time required to obtain test results, sampling for the test on exit was assumed to occur the day before the quarantine was completed. Cells that share a background color in common indicate equivalent durations of quarantine associated with each of the testing strategies.

**Figure S13:**
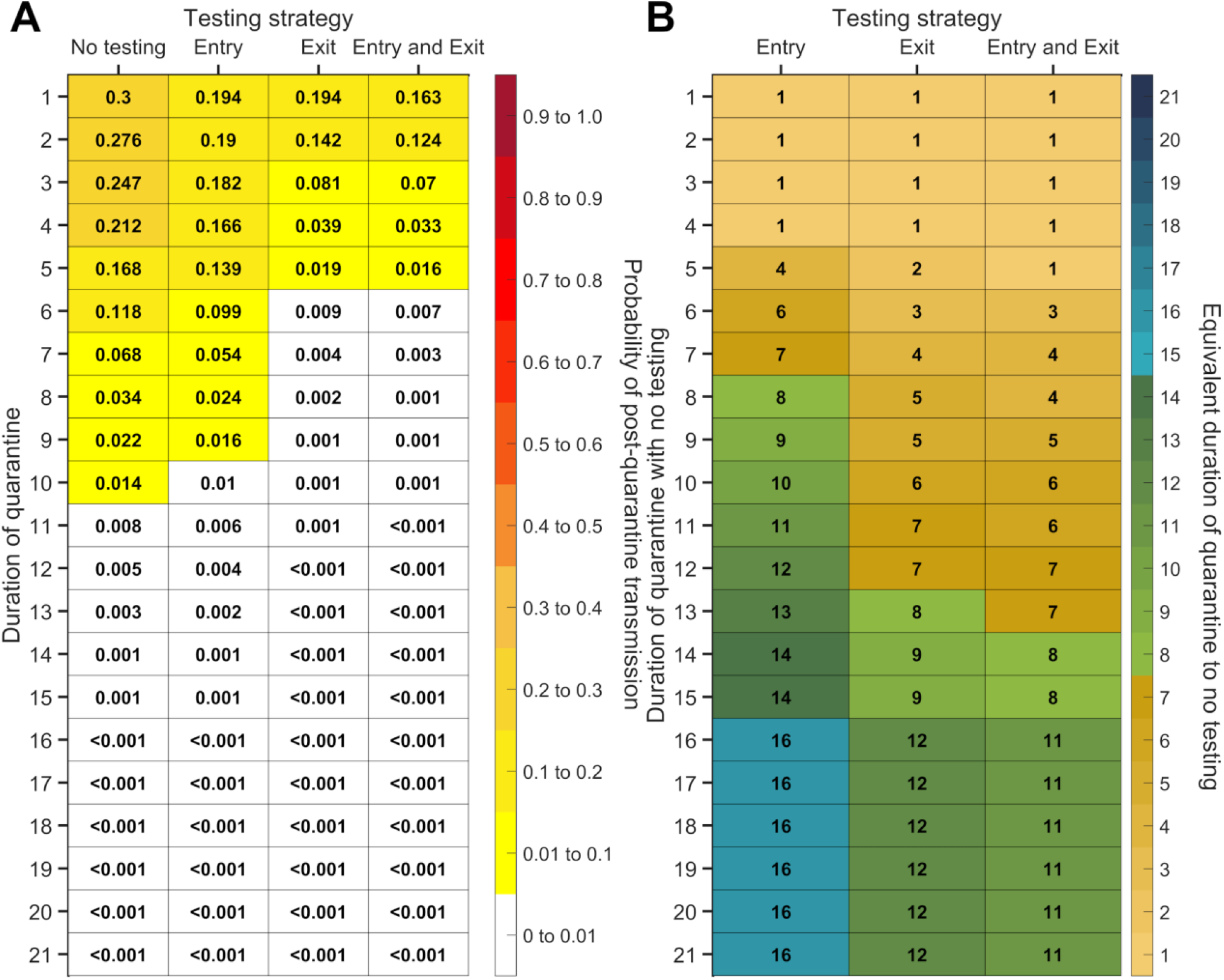
For durations of quarantine from 1–21 days, when a symptomatic individual enters quarantine uniformly within the incubation period and asymptomatic individuals enter uniformly across the disease time course, with an incubation period of 8.29 days, a latent period of 1.9 days, with 30.8% of infections being asymptomatic, and perfect self-isolation of symptomatic infections, (**A**) the probability of post-quarantine transmission (probability of one or more post-quarantine infections) with no testing, when tested upon entry to quarantine, when tested on exit from quarantine, and when tested on entry and exit from quarantine, and (**B**) the durations of quarantine with testing on entry, testing on exit, and testing on entry and exit that perform just as well or better than a quarantine with no testing. Because of the time required to obtain test results, sampling for the test on exit is assumed to occur the day before the quarantine is complete. Cells that share a background color in common indicate equivalent durations of quarantine associated with each of the testing strategies.

**Figure S14:**
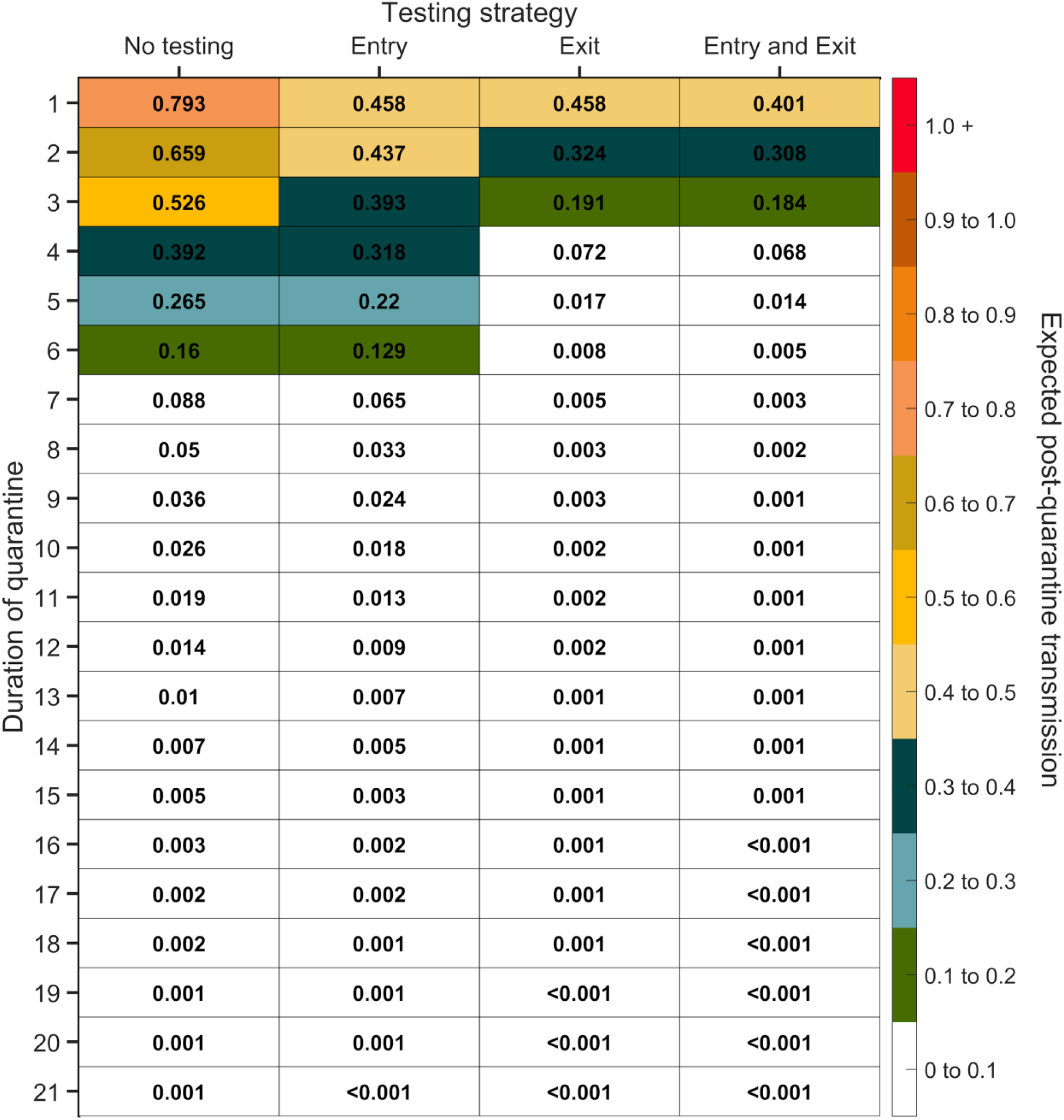
Expected post-quarantine infections for durations of quarantine of 1–21 days, with an incubation period of 8.29 days, a latent period of 3.9 days, 30.8% of infections being asymptomatic, perfect self-isolation of symptomatic infections when symptomatic, uniform entry within the incubation period by symptomatic cases, and uniform entry across the disease time course for asymptomatic cases, with no testing, testing on entry, testing on exit, and testing on entry and exit. Because of the time required to obtain test results, sampling for the test on exit was assumed to occur the day before the quarantine was completed. Cells that share a background color in common indicate equivalent durations of quarantine associated with each of the testing strategies.

**Figure S15:**
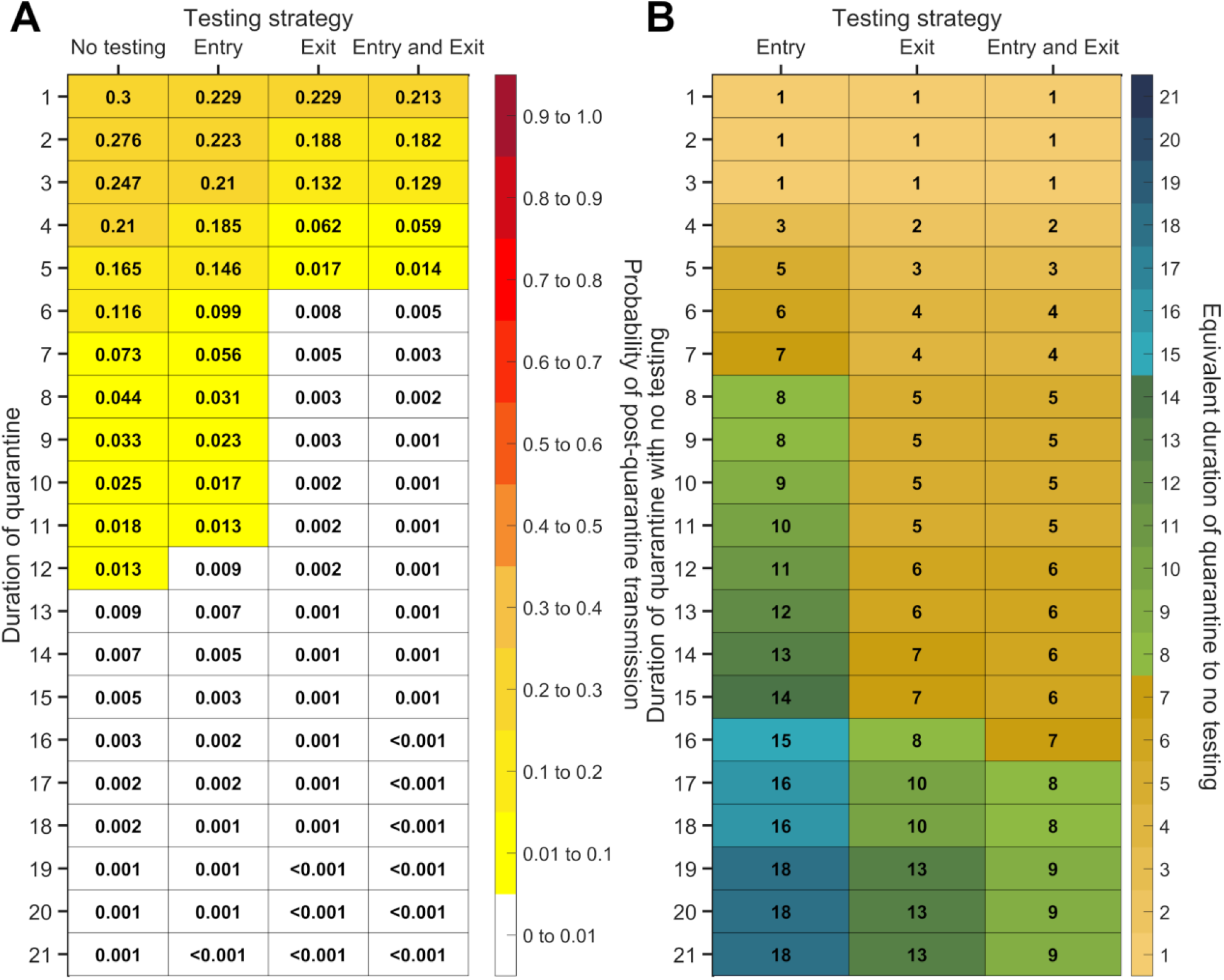
For durations of quarantine from 1–21 days, when a symptomatic individual enters quarantine uniformly within the incubation period and asymptomatic individuals enter uniformly across the disease time course, with an incubation period of 8.29 days, a latent period of 3.9 days, with 30.8% of infections being asymptomatic, and perfect self-isolation of symptomatic infections, (**A**) the probability of post-quarantine transmission (probability of one or more post-quarantine infections) with no testing, when tested upon entry to quarantine, when tested on exit from quarantine, and when tested on entry and exit from quarantine, and (**B**) the durations of quarantine with testing on entry, testing on exit, and testing on entry and exit that perform just as well or better than a quarantine with no testing. Because of the time required to obtain test results, sampling for the test on exit is assumed to occur the day before the quarantine is complete. Cells that share a background color in common indicate equivalent durations of quarantine associated with each of the testing strategies.

**Figure S16:**
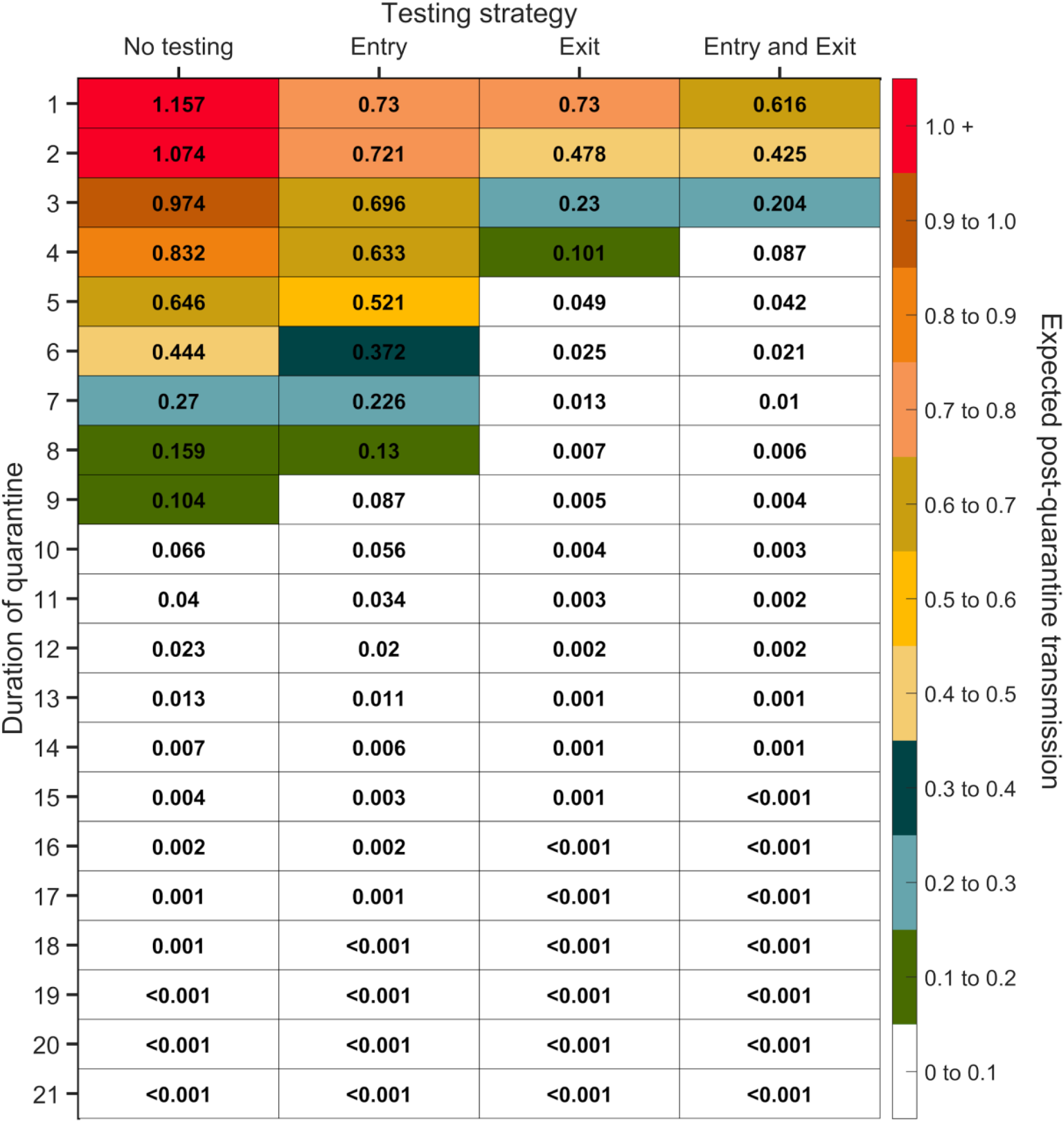
Expected post-quarantine infections for durations of quarantine of 1–21 days, with an incubation period of 8.29 days, a latent period of 1.9 days, 30.8% of infections being asymptomatic, perfect self-isolation of symptomatic infections when symptomatic, and entry through contact tracing, with no testing, testing on entry, testing on exit, and testing on entry and exit. Because of the time required to obtain test results, sampling for the test on exit was assumed to occur the day before the quarantine was completed. Cells that share a background color in common indicate equivalent durations of quarantine associated with each of the testing strategies.

**Figure S17:**
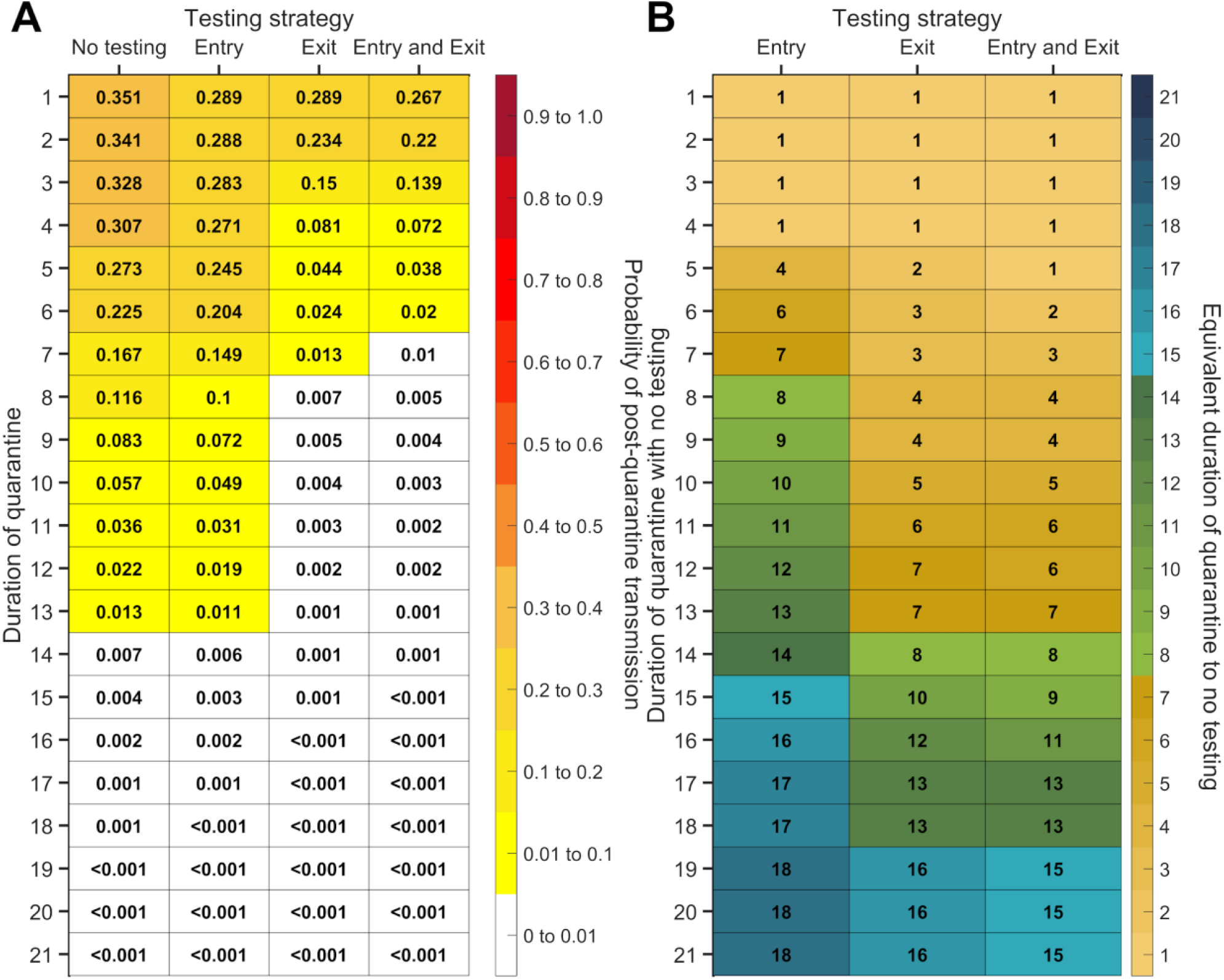
For durations of quarantine from 1–21 days, when an individual enters quarantine through contact tracing, with an incubation period of 8.29 days, a latent period of 1.9 days, with 30.8% of infections being asymptomatic, and perfect self-isolation of symptomatic infections, (**A**) the probability of post-quarantine transmission (probability of one or more post-quarantine infections) with no testing, when tested upon entry to quarantine, when tested on exit from quarantine, and when tested on entry and exit from quarantine, and (**B**) the durations of quarantine with testing on entry, testing on exit, and testing on entry and exit that perform just as well or better than a quarantine with no testing. Because of the time required to obtain test results, sampling for the test on exit is assumed to occur the day before the quarantine is complete. Cells that share a background color in common indicate equivalent durations of quarantine associated with each of the testing strategies.

**Figure S18:**
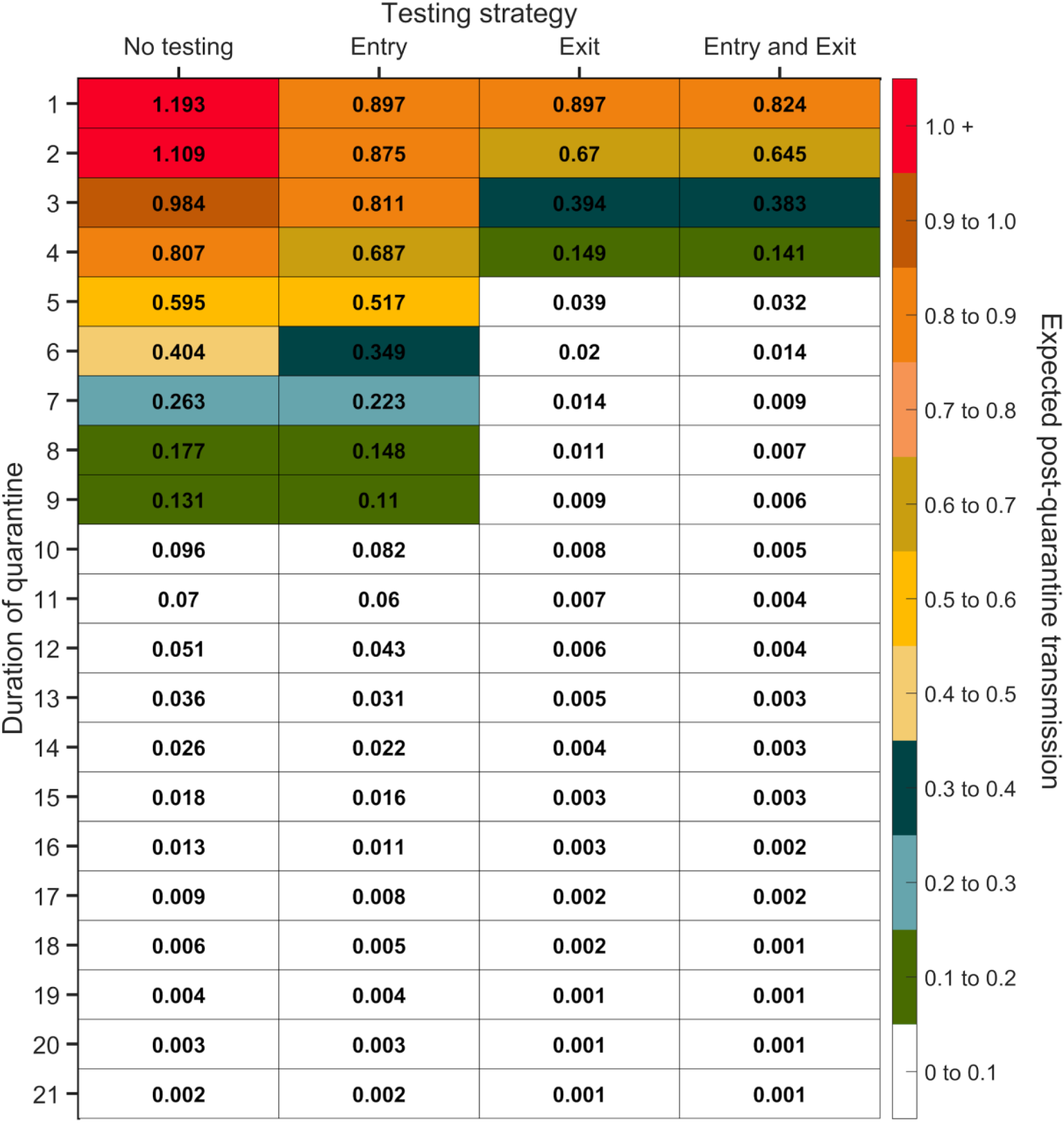
Expected post-quarantine infections for durations of quarantine of 1–21 days, with an incubation period of 8.29 days, a latent period of 3.9 days, 30.8% of infections being asymptomatic, perfect self-isolation of symptomatic infections when symptomatic, and entry through contact tracing, with no testing, testing on entry, testing on exit, and testing on entry and exit. Because of the time required to obtain test results, sampling for the test on exit was assumed to occur the day before the quarantine was completed. Cells that share a background color in common indicate equivalent durations of quarantine associated with each of the testing strategies.

**Figure S19:**
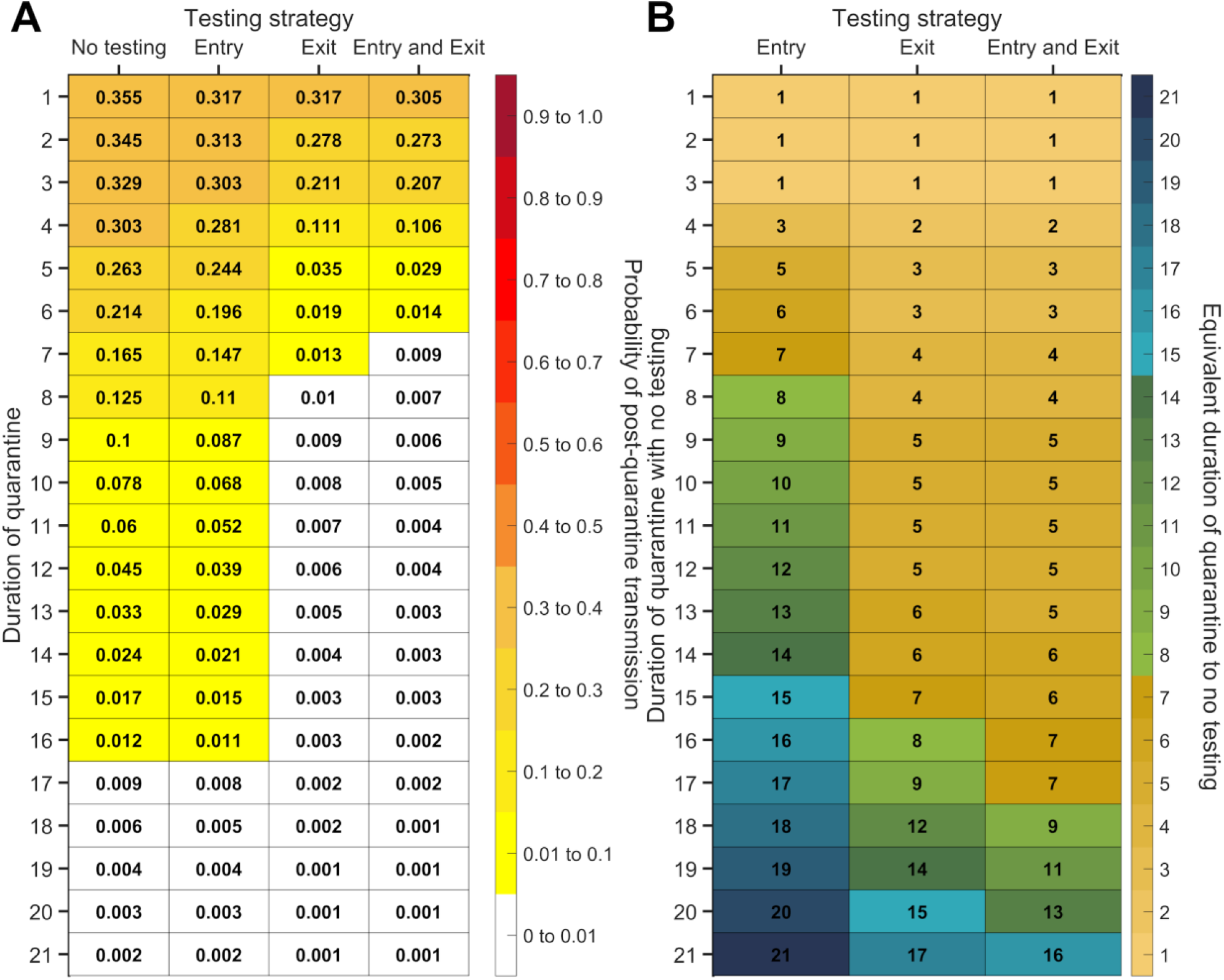
For durations of quarantine from 1–21 days, when an individual enters quarantine through contact tracing, with an incubation period of 8.29 days, a latent period of 3.9 days, with 30.8% of infections being asymptomatic, and perfect self-isolation of symptomatic infections, (**A**) the probability of post-quarantine transmission (probability of one or more post-quarantine infections) with no testing, when tested upon entry to quarantine, when tested on exit from quarantine, and when tested on entry and exit from quarantine, and (**B**) the durations of quarantine with testing on entry, testing on exit, and testing on entry and exit that perform just as well or better than a quarantine with no testing. Because of the time required to obtain test results, sampling for the test on exit is assumed to occur the day before the quarantine is complete. Cells that share a background color in common indicate equivalent durations of quarantine associated with each of the testing strategies.

**Figure S20:**
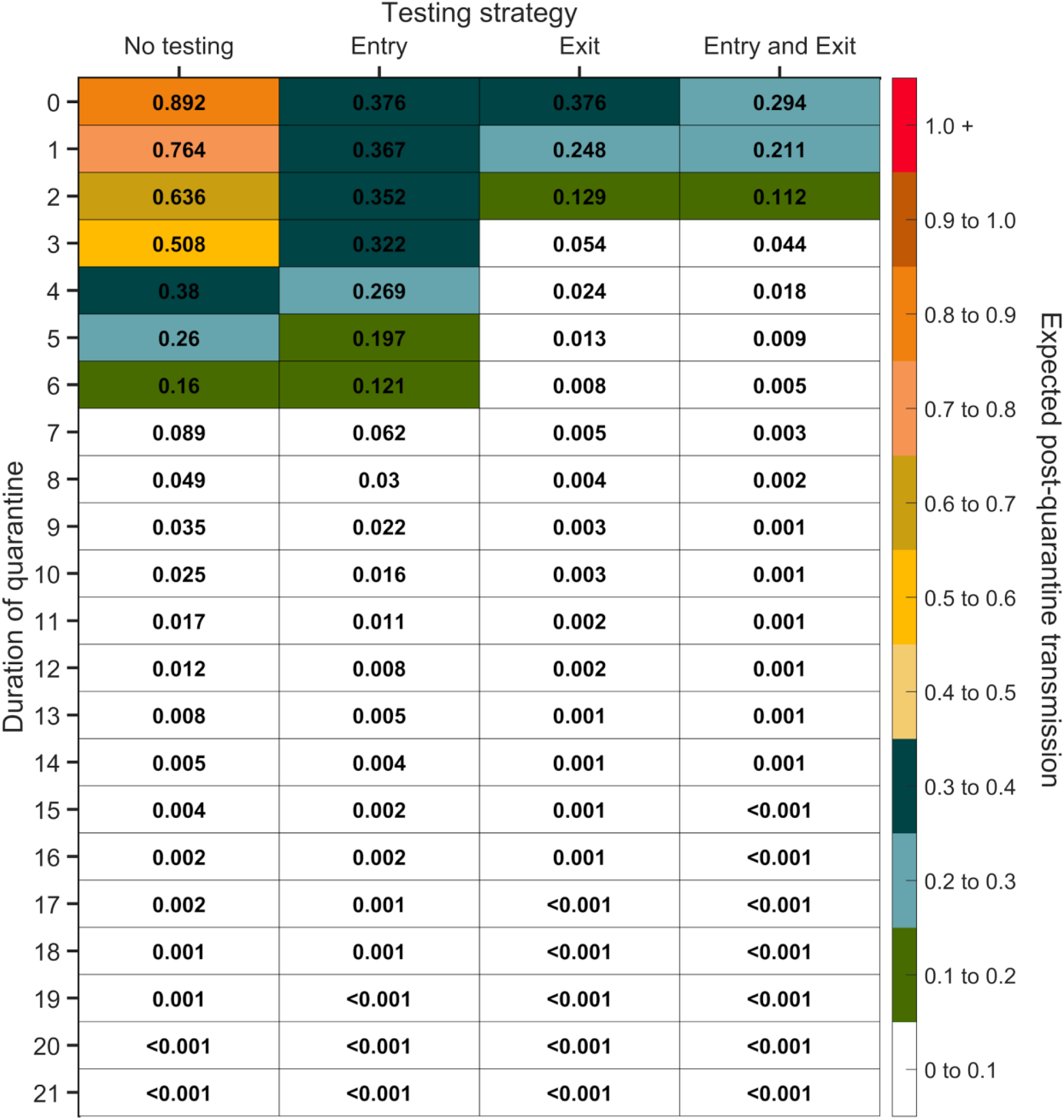
Expected post-quarantine infections for durations of quarantine of 0–21 days, with an incubation period of 8.29 days, a latent period of 2.9 days, 30.8% of infections being asymptomatic, perfect self-isolation of symptomatic infections when symptomatic, uniform entry within the incubation period by symptomatic cases, and uniform entry across the disease time course for asymptomatic cases, with no testing, testing on entry, testing on exit, and testing on entry and exit. Testing on exit is assumed to occur on the last day of quarantine (i.e. there is negligible delay in obtaining the test result). Cells that share a background color in common indicate equivalent durations of quarantine associated with each of the testing strategies.

**Figure S21:**
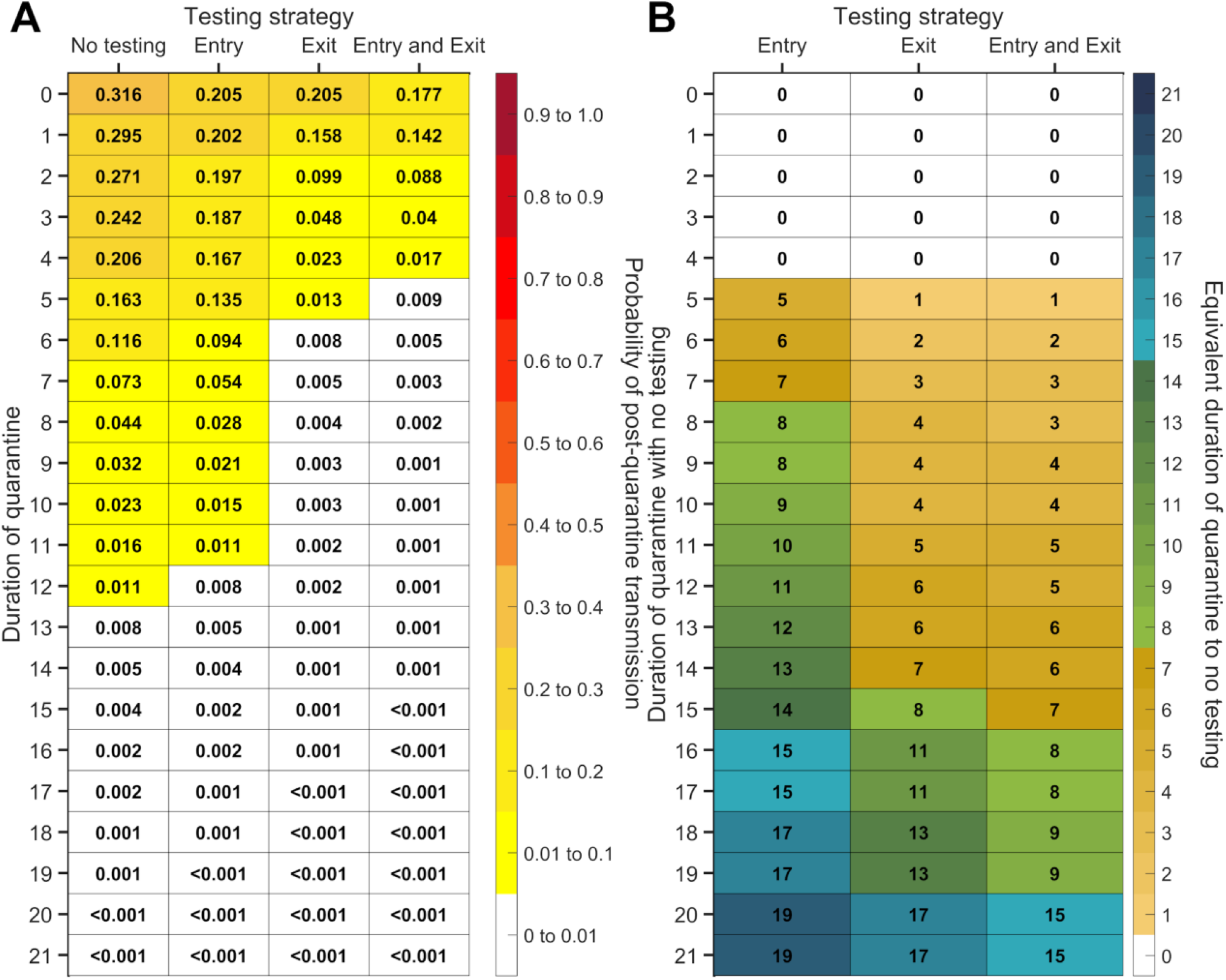
For durations of quarantine from 0–21 days, when a symptomatic individual enters quarantine uniformly within the incubation period and asymptomatic individuals enter uniformly across the disease time course, with an incubation period of 8.29 days, a latent period of 2.9 days, with 30.8% of infections being asymptomatic, and perfect self-isolation of symptomatic infections, (**A**) the probability of post-quarantine transmission (probability of one or more post-quarantine infections) with no testing, when tested upon entry to quarantine, when tested on exit from quarantine, and when tested on entry and exit from quarantine, and (**B**) the durations of quarantine with testing on entry, testing on exit, and testing on entry and exit that perform just as well or better than a quarantine with no testing. Testing on exit is assumed to occur on the last day of quarantine (i.e. there is a negligible delay in obtaining the test results). Cells that share a background color in common indicate equivalent durations of quarantine associated with each of the testing strategies.

**Figure S22:**
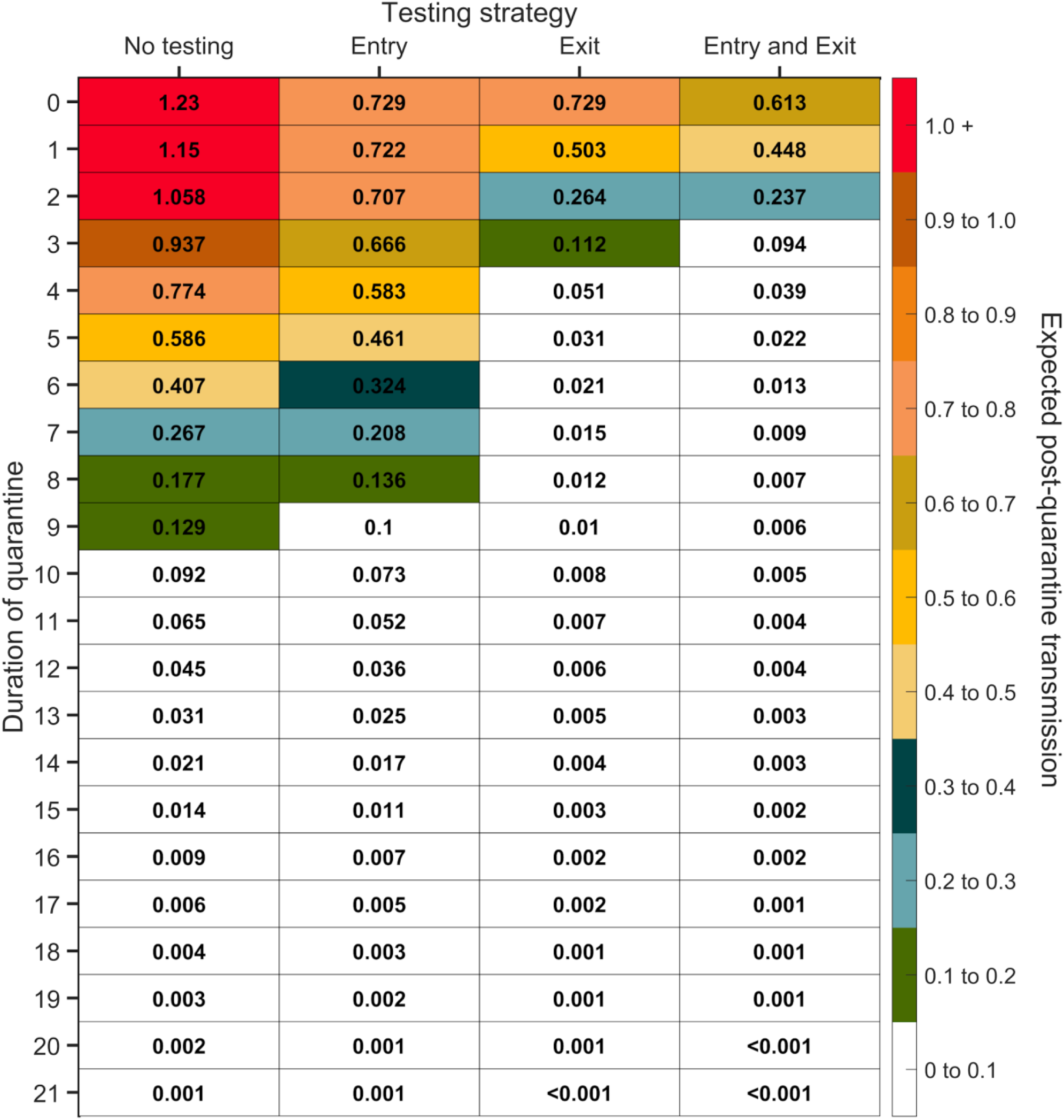
Expected post-quarantine infections for durations of quarantine of 0–21 days, with an incubation period of 8.29 days, a latent period of 2.9 days, 30.8% of infections being asymptomatic, perfect self-isolation of symptomatic infections when symptomatic, and entry through contact tracing, with no testing, testing on entry, testing on exit, and testing on entry and exit. Testing on exit is assumed to occur on the last day of quarantine (i.e. there is negligible delay in obtaining the test result). Cells that share a background color in common indicate equivalent durations of quarantine associated with each of the testing strategies.

**Figure S23:**
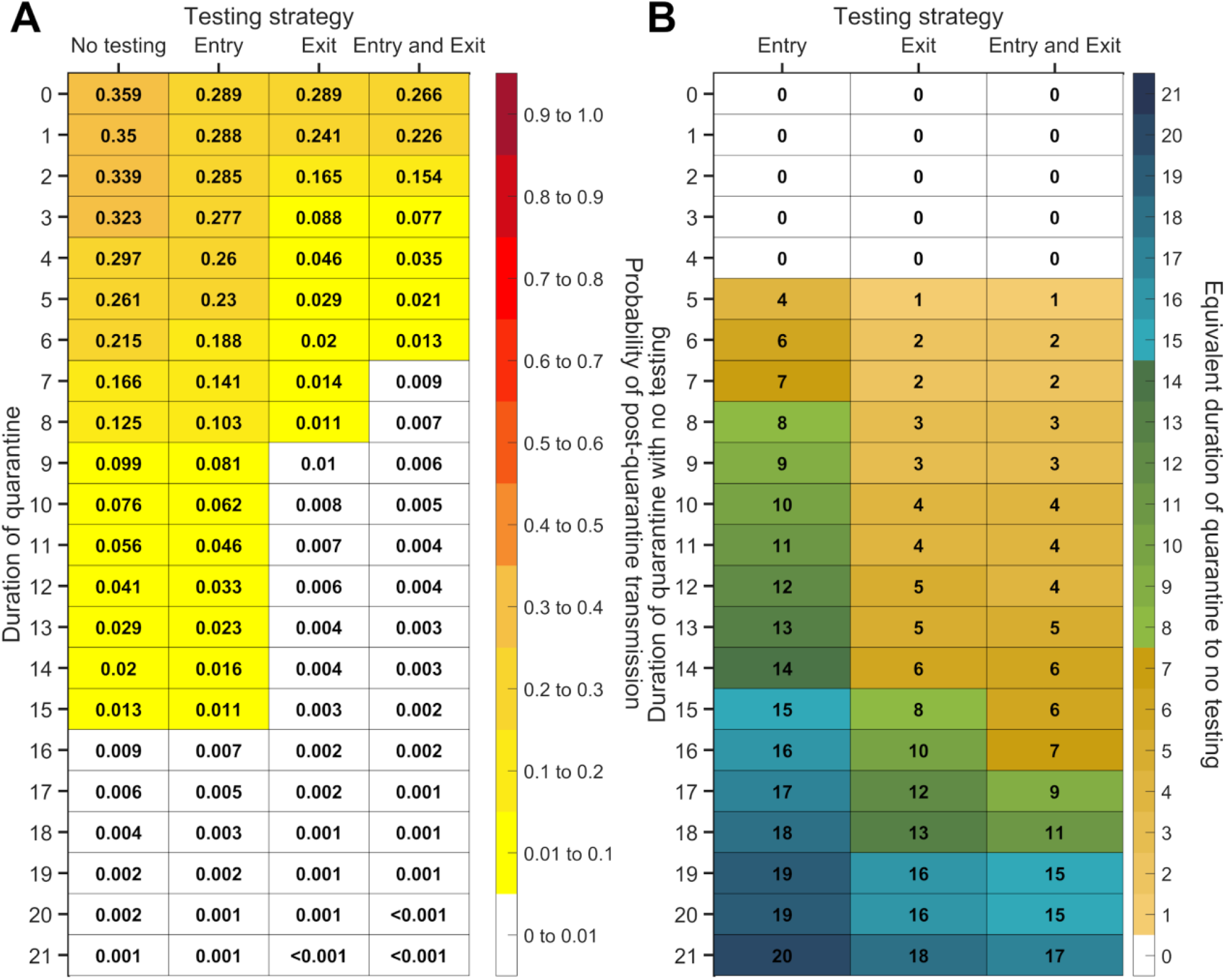
For durations of quarantine from 0–21 days, when an individual enters quarantine through contact tracing, with an incubation period of 8.29 days, a latent period of 2.9 days, with 30.8% of infections being asymptomatic, and perfect self-isolation of symptomatic infections, (**A**) the probability of post-quarantine transmission (probability of one or more post-quarantine infections) with no testing, when tested upon entry to quarantine, when tested on exit from quarantine, and when tested on entry and exit from quarantine, and (**B**) the durations of quarantine with testing on entry, testing on exit, and testing on entry and exit that perform just as well or better than a quarantine with no testing. Testing on exit is assumed to occur on the last day of quarantine (i.e. there is a negligible delay in obtaining the test result). Cells that share a background color in common indicate equivalent durations of quarantine associated with each of the testing strategies.

**Figure S24:**
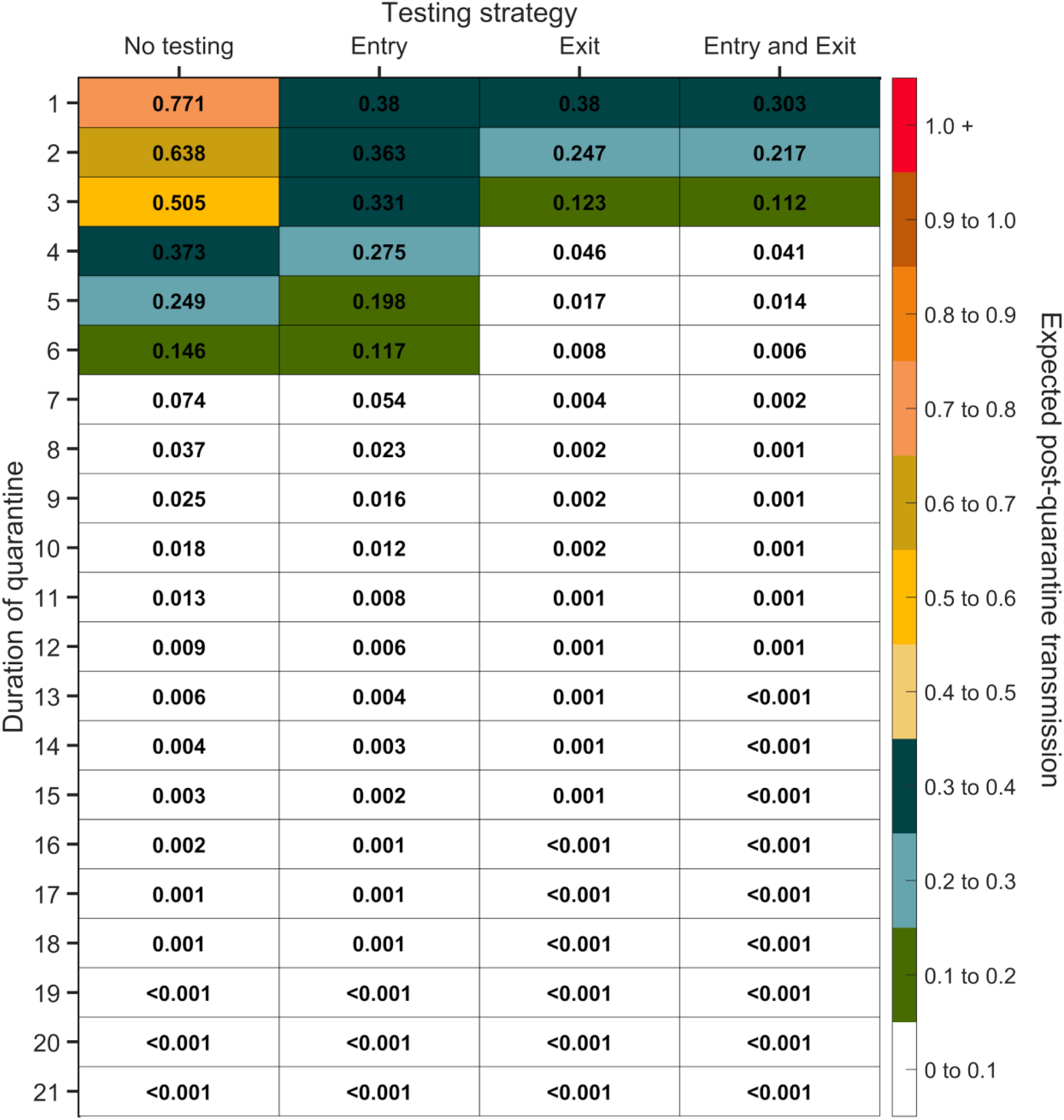
Expected post-quarantine infections for durations of quarantine of 1–21 days, with an incubation period of 8.29 days, a latent period of 2.9 days, 22.6% of infections being asymptomatic, perfect self-isolation of symptomatic infections when symptomatic, uniform entry within the incubation period by symptomatic cases, and uniform entry across the disease time course for asymptomatic cases, with no testing, testing on entry, testing on exit, and testing on entry and exit. Because of the time required to obtain test results, sampling for the test on exit was assumed to occur the day before the quarantine was completed. Cells that share a background color in common indicate equivalent durations of quarantine associated with each of the testing strategies.

**Figure S25:**
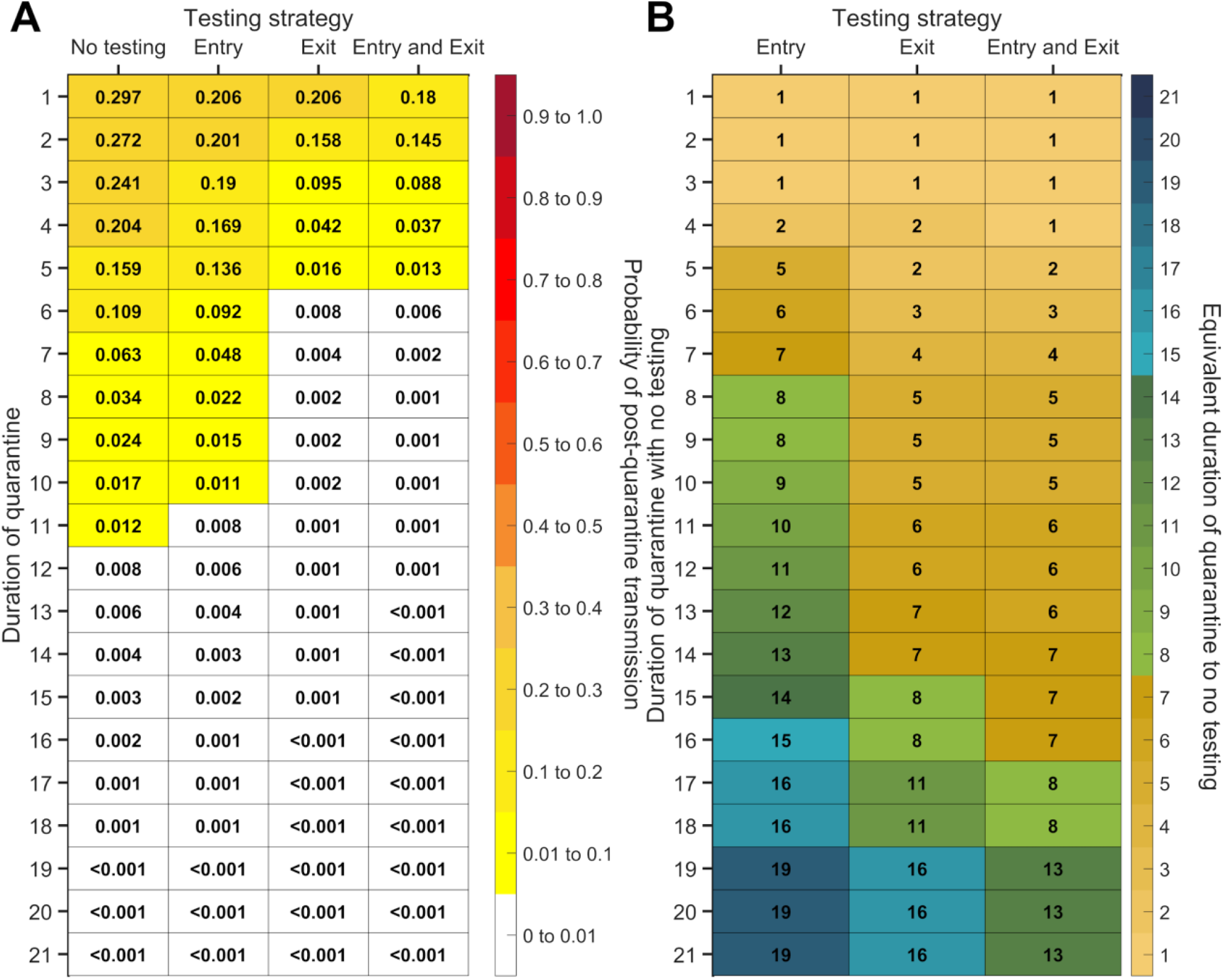
For durations of quarantine from 1–21 days, when a symptomatic individual enters quarantine uniformly within the incubation period and asymptomatic individuals enter uniformly across the disease time course, with an incubation period of 8.29 days, a latent period of 2.9 days, with 22.6% of infections being asymptomatic, and perfect self-isolation of symptomatic infections, (**A**) the probability of post-quarantine transmission (probability of one or more post-quarantine infections) with no testing, when tested upon entry to quarantine, when tested on exit from quarantine, and when tested on entry and exit from quarantine, and (**B**) the durations of quarantine with testing on entry, testing on exit, and testing on entry and exit that perform just as well or better than a quarantine with no testing. Because of the time required to obtain test results, sampling for the test on exit is assumed to occur the day before the quarantine is complete. Cells that share a background color in common indicate equivalent durations of quarantine associated with each of the testing strategies.

**Figure S26:**
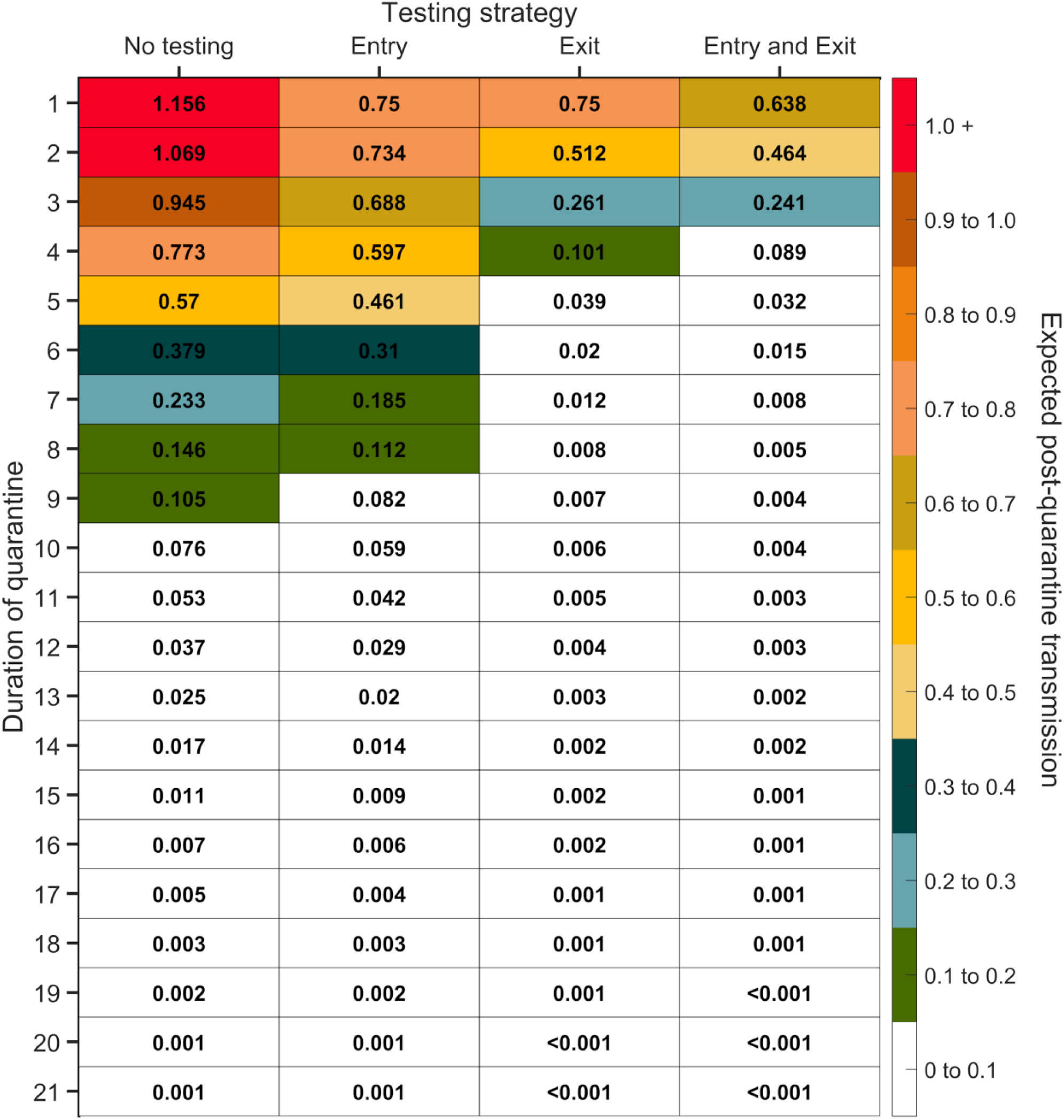
Expected post-quarantine infections for durations of quarantine of 1–21 days, with an incubation period of 8.29 days, a latent period of 2.9 days, 22.6% of infections being asymptomatic, perfect self-isolation of symptomatic infections when symptomatic, and entry through contact tracing, with no testing, testing on entry, testing on exit, and testing on entry and exit. Because of the time required to obtain test results, sampling for the test on exit was assumed to occur the day before the quarantine was completed. Cells that share a background color in common indicate equivalent durations of quarantine associated with each of the testing strategies.

**Figure S27:**
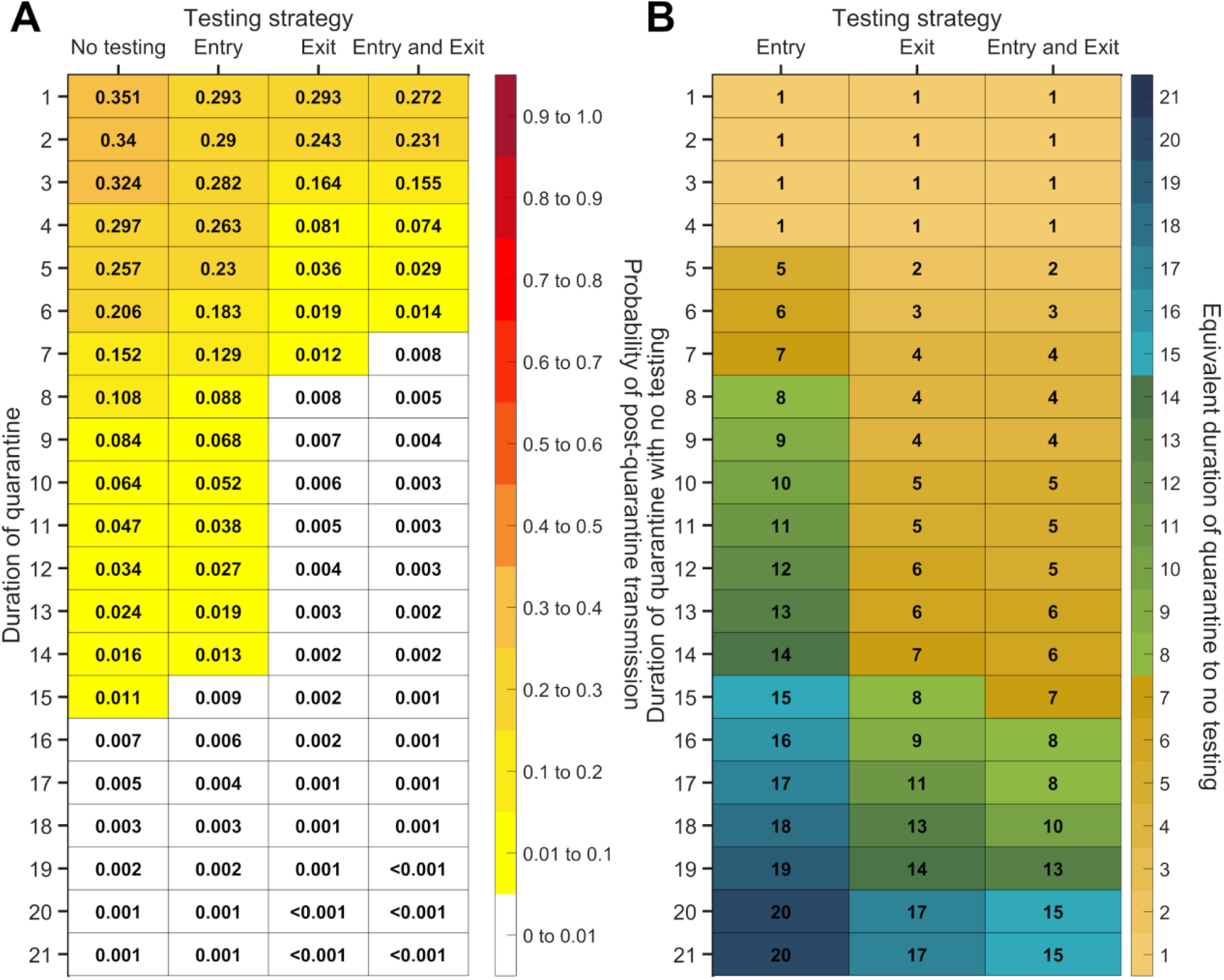
For durations of quarantine from 1–21 days, when an individual enters quarantine through contact tracing, with an incubation period of 8.29 days, a latent period of 2.9 days, with 22.6% of infections being asymptomatic, and perfect self-isolation of symptomatic infections, (**A**) the probability of post-quarantine transmission (probability of one or more post-quarantine infections) with no testing, when tested upon entry to quarantine, when tested on exit from quarantine, and when tested on entry and exit from quarantine, and (**B**) the durations of quarantine with testing on entry, testing on exit, and testing on entry and exit that perform just as well or better than a quarantine with no testing. Because of the time required to obtain test results, sampling for the test on exit is assumed to occur the day before the quarantine is complete. Cells that share a background color in common indicate equivalent durations of quarantine associated with each of the testing strategies.

**Figure S28:**
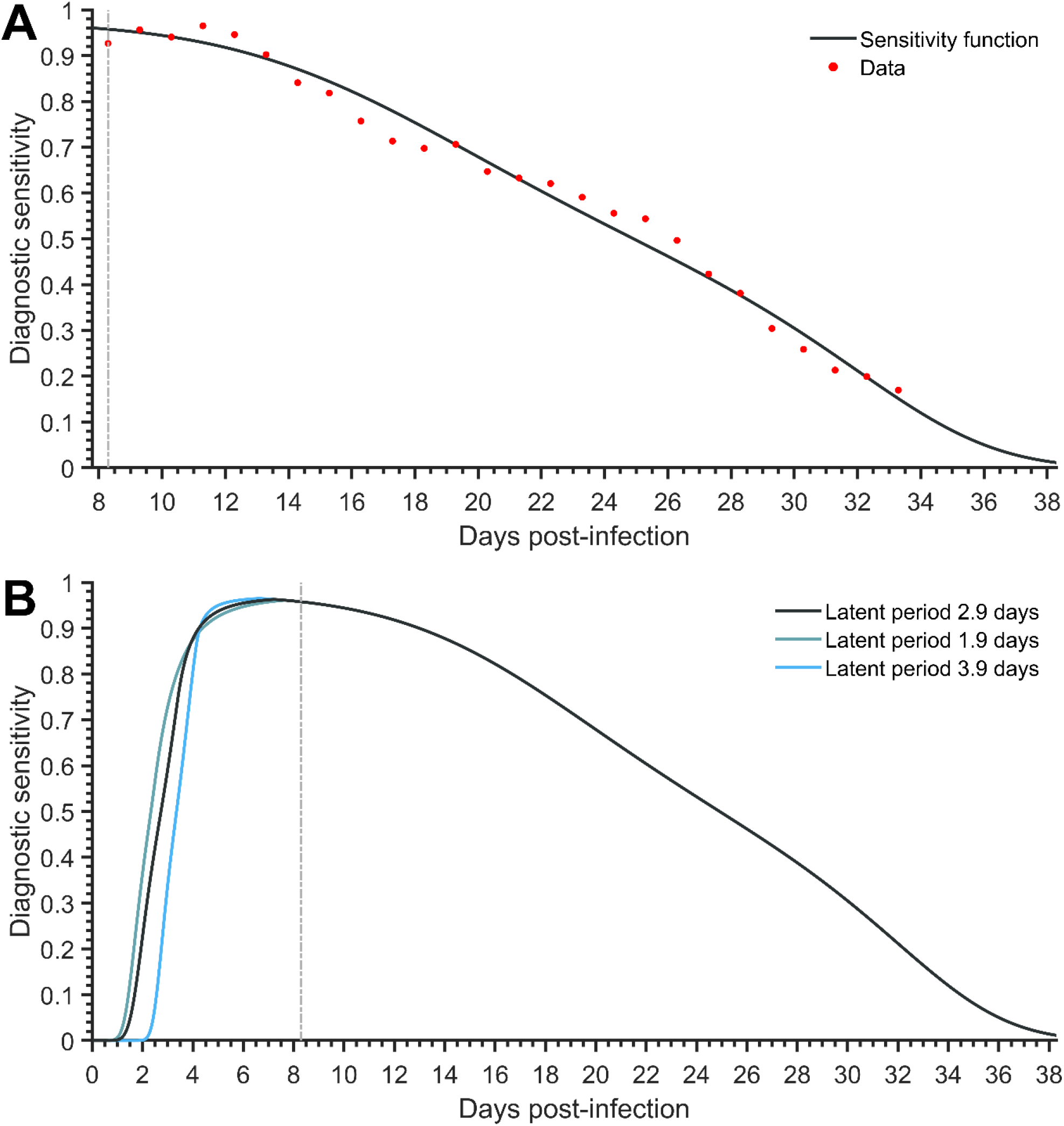
For an incubation period of 8.29 days, the diagnostic sensitivity of the RT-PCR test over the time course of disease (**A**) determined using a logistic regression model (black line) fit to the empirical data of SARS CoV-2 test results from Miller et al 6(red dots) through minimization of least squares and AIC model selection, and (**B**) specifying latent periods of 2.9 days (black), 1.9 days (green), and 3.9 days (blue).

**Figure S29:**
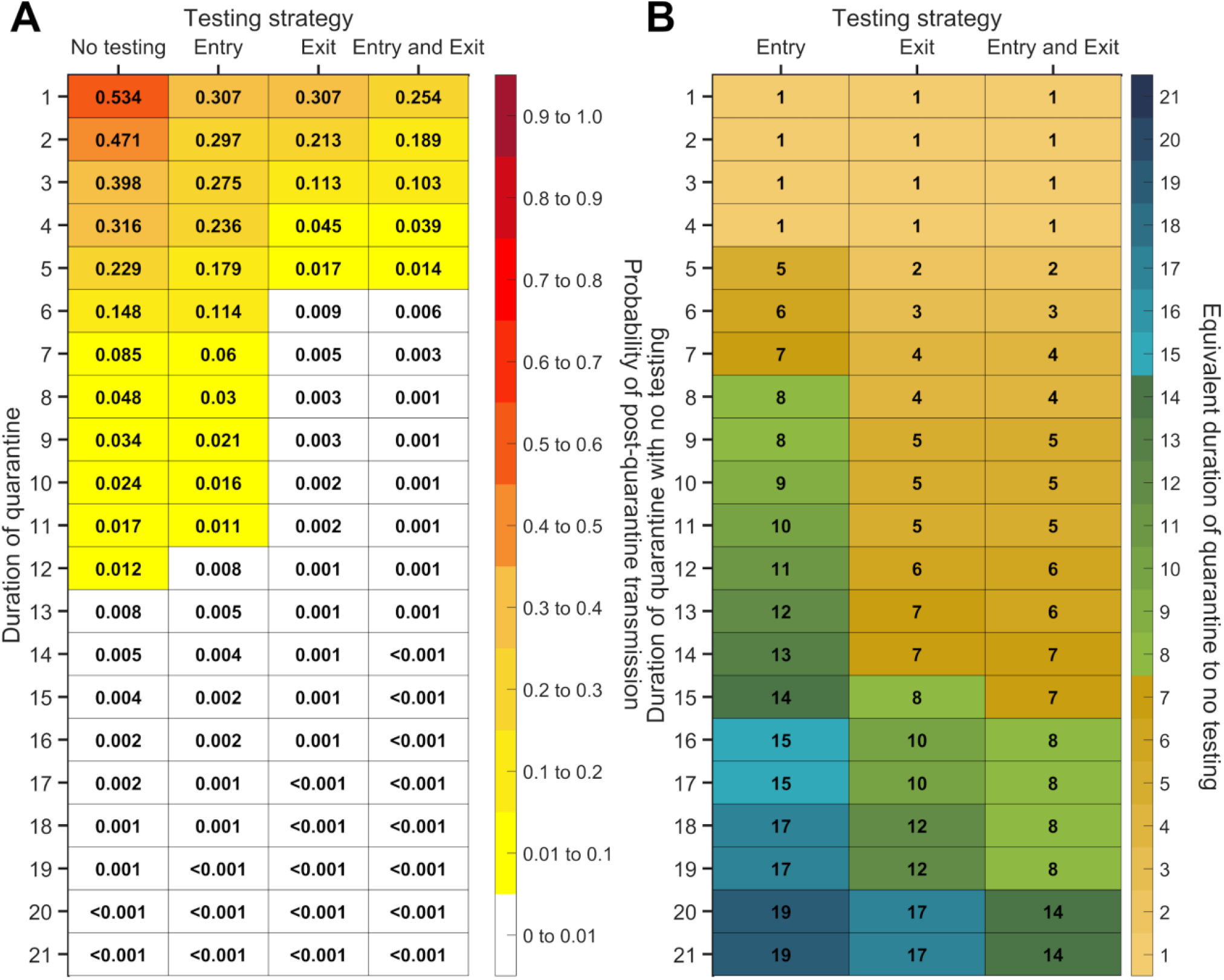
For durations of quarantine from 1–21 days, when a symptomatic individual enters quarantine uniformly within the incubation period and asymptomatic individuals enter uniformly across the disease time course, with an incubation period of 8.29 days, a latent period of 2.9 days, with 30.8% of infections being asymptomatic, and perfect self-isolation of symptomatic infections, (**A**) the probability of post-quarantine transmission (probability of one or more post-quarantine infections), assuming infections are Poisson distributed, with no testing, when tested upon entry to quarantine, when tested on exit from quarantine, and when tested on entry and exit from quarantine, and (**B**) the durations of quarantine with testing on entry, testing on exit, and testing on entry and exit that perform just as well or better than a quarantine with no testing. Because of the time required to obtain test results, sampling for the test on exit is assumed to occur the day before the quarantine is complete. Cells that share a background color in common indicate equivalent durations of quarantine associated with each of the testing strategies.

**Figure S30:**
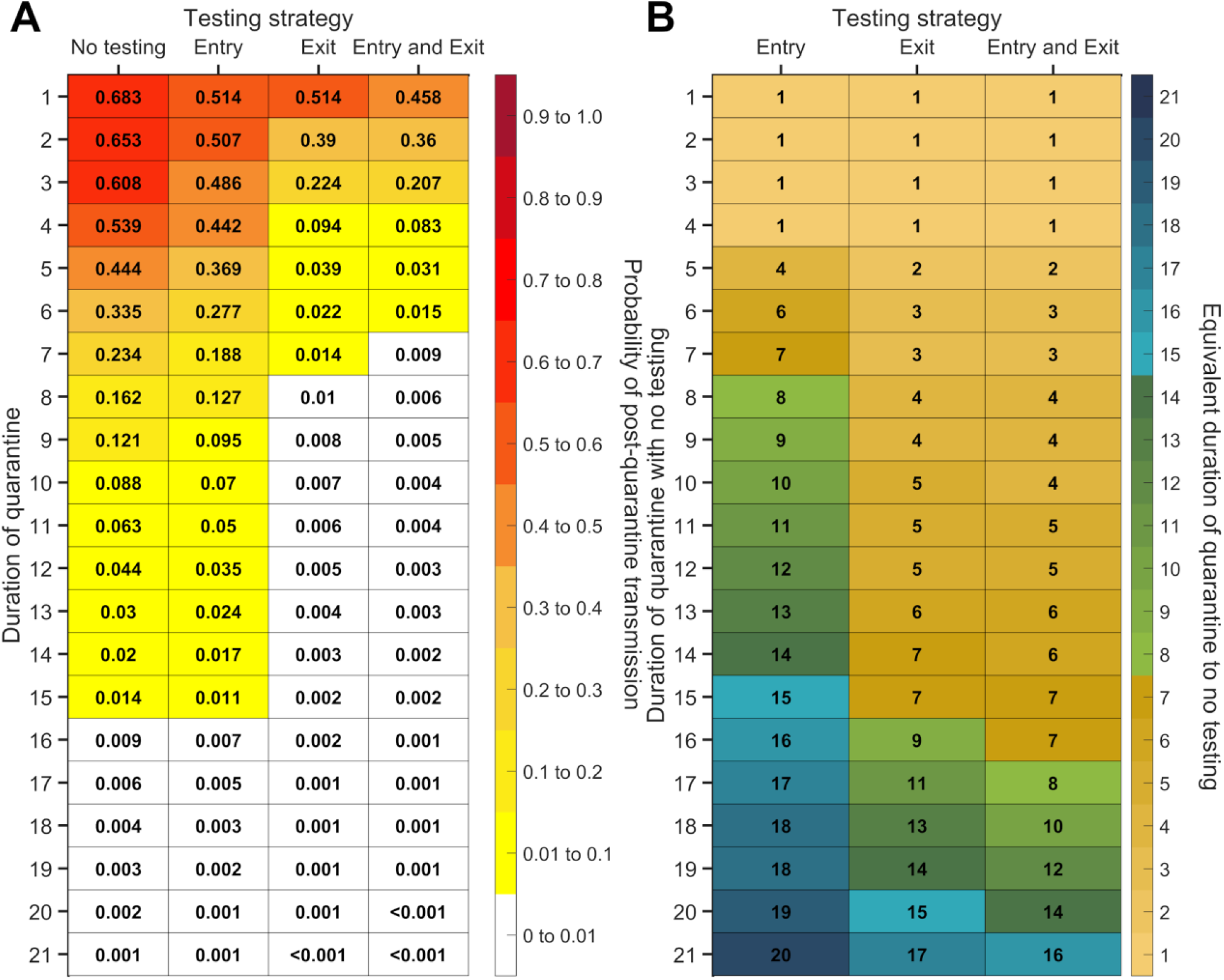
For durations of quarantine from 1–21 days, when an individual enters quarantine through contact tracing, with an incubation period of 8.29 days, a latent period of 2.9 days, with 30.8% of infections being asymptomatic, and perfect self-isolation of symptomatic infections, (**A**) the probability of post-quarantine transmission (probability of one or more post-quarantine infections), assuming infections are Poisson distributed, with no testing, when tested upon entry to quarantine, when tested on exit from quarantine, and when tested on entry and exit from quarantine, and (**B**) the durations of quarantine with testing on entry, testing on exit, and testing on entry and exit that perform just as well or better than a quarantine with no testing. Because of the time required to obtain test results, sampling for the test on exit is assumed to occur the day before the quarantine is complete. Cells that share a background color in common indicate equivalent durations of quarantine associated with each of the testing strategies.

**Figure S31:**
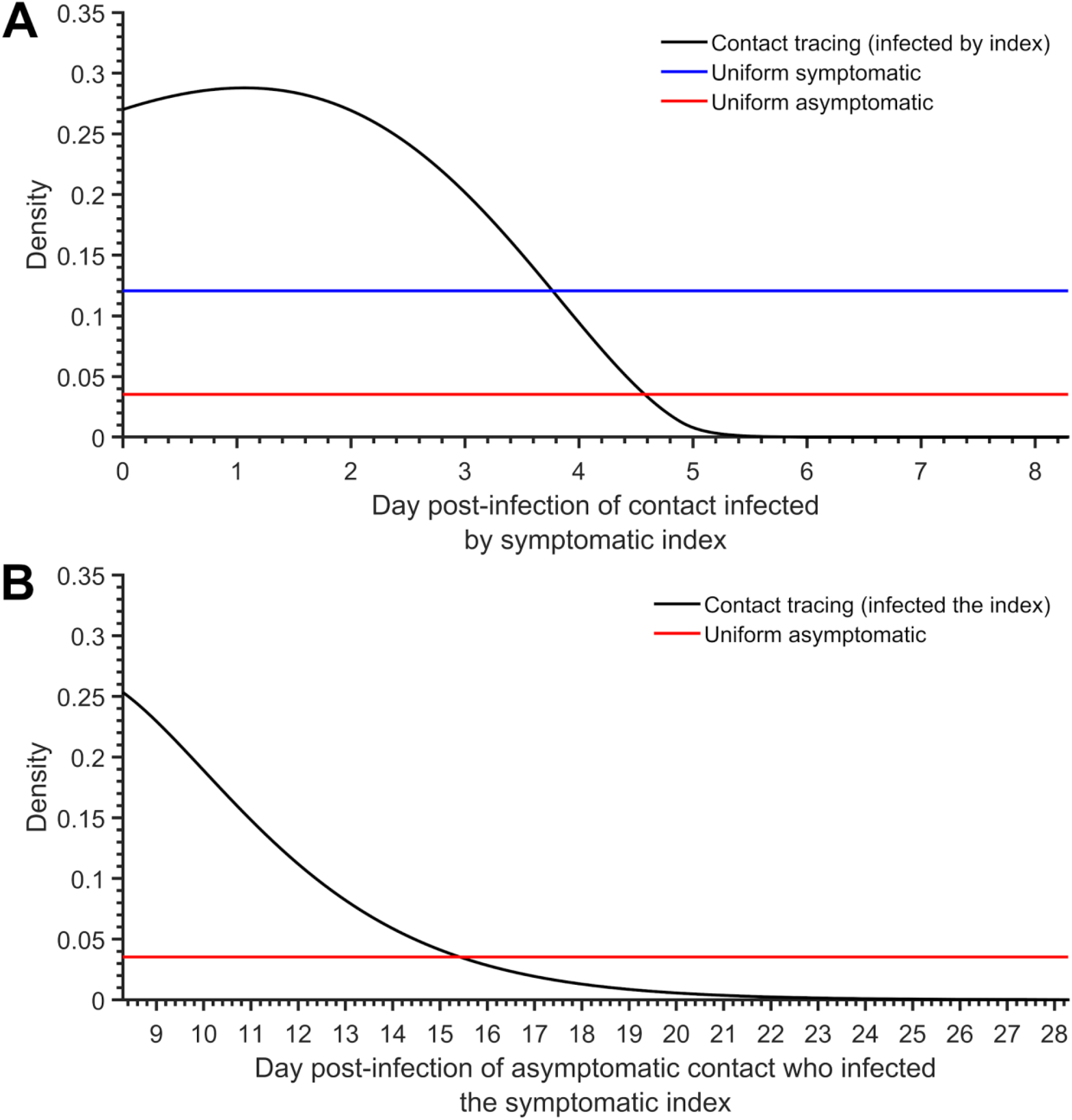
Probability density functions for when during the disease time course cases enter quarantine, including (**A**) the day of disease time course in which a contact infected by an index case enters quarantine (black) compared to the uniform entry into quarantine of a case to exhibit symptoms (blue) and an asymptomatic case (red), and (**B**) the day of disease time course in which the asymptomatic contact that infected the index case enters quarantine (black) compared to the uniform entry into quarantine of an asymptomatic case (red).

## Notes

### Competing Interest Statement

JTP, GK,BS,RHM,SMM, APG received funding from the company who collected and provided data

### Author Declarations

IRB Approval from the Office of Research Ethics (ORE) at York University, Toronto ON, Canada Certificate #: 2020-323 (Approval Document can be provided if requested)

## References

1. Nicola, M. et al.. The socio-economic implications of the coronavirus pandemic (COVID-19): A review. Int. J. Surg. 78, 185–193 (2020).

2. Martin, A., Markhvida, M., Hallegatte, S. & Walsh, B. Socio-Economic Impacts of COVID-19 on Household Consumption and Poverty. Econ Disaster Clim Chang 1–27 (2020).

3. Lee, J. C., Mervosh, S., Avila, Y., Harvey, B. & Matthews, A. L. See How All 50 States Are Reopening (and Closing Again). The New York Times (2020).

4. Aleta, A. et al. Modelling the impact of testing, contact tracing and household quarantine on second waves of COVID-19. Nature Human Behaviour 4, 964–971 (2020).

5. CDC. When to Quarantine. https://www.cdc.gov/coronavirus/2019-ncov/if-you-are-sick/quarantine.html (2020).

6. Brooks, S. K. et al. The psychological impact of quarantine and how to reduce it: rapid review of the evidence. Lancet 395, 912–920 (2020).

7. Mattioli, A. V., Puviani, M. B., Nasi, M. & Farinetti, A. COVID-19 pandemic: the effects of quarantine on cardiovascular risk. Eur. J. Clin. Nutr. 74, 852–855 (2020).

8. Moghadas, S. M. et al. The implications of silent transmission for the control of COVID-19 outbreaks. Proc. Natl. Acad. Sci. U. S. A. (2020) doi:10.1073/pnas.2008373117.

9. Gondim, J. A. M. & Machado, L. Optimal quarantine strategies for the COVID-19 pandemic in a population with a discrete age structure. Chaos, Solitons & Fractals vol. 140 110166 (2020).

10. Cui, Q. et al. Dynamic variations of the COVID-19 disease at different quarantine strategies in Wuhan and mainland China. J. Infect. Public Health 13, 849–855 (2020).

11. Hou, C. et al. The effectiveness of quarantine of Wuhan city against the Corona Virus Disease 2019 (COVID-19): A well-mixed SEIR model analysis. J. Med. Virol. 92, 841–848 (2020).

12. Quarantine and testing strategies in contact tracing for SARS-CoV-2. https://cmmid.github.io/topics/covid19/quar-test-contact-tracing.html (2020).

13. Grassly, N. C. et al. Comparison of molecular testing strategies for COVID-19 control: a mathematical modelling study. Lancet Infect. Dis. (2020) doi:10.1016/S1473-3099(20)30630-7.

14. Kucirka, L. M., Lauer, S. A., Laeyendecker, O., Boon, D. & Lessler, J. Variation in False-Negative Rate of Reverse Transcriptase Polymerase Chain Reaction–Based SARS-CoV-2 Tests by Time Since Exposure. Annals of Internal Medicine vol. 173 262–267 (2020).

15. Jung, J. et al. The Importance of Mandatory COVID-19 Diagnostic Testing Prior to Release from Quarantine. J. Korean Med. Sci. 35, (2020).

16. Australian Government Department of Health. Coronavirus (COVID-19) advice for international travellers. https://www.health.gov.au/news/health-alerts/novel-coronavirus-2019-ncov-health-alert/coronavirus-covid-19-restrictions/coronavirus-covid-19-advice-for-international-travellers (2020).

17. He, X. et al. Author Correction: Temporal dynamics in viral shedding and transmissibility of COVID-19. Nat. Med. (2020) doi:10.1038/s41591-020-1016-z.

18. Qin, J. et al. Estimation of incubation period distribution of COVID-19 using disease onset forward time: A novel cross-sectional and forward follow-up study. Sci Adv 6, eabc1202 (2020).

19. Miller, T. E. et al. Clinical sensitivity and interpretation of PCR and serological COVID-19 diagnostics for patients presenting to the hospital. FASEB J. (2020) doi:10.1096/fj.202001700RR.

20. Nishiura, H. et al. Estimation of the asymptomatic ratio of novel coronavirus infections (COVID-19). Int. J. Infect. Dis. 94, 154–155 (2020).

21. Buitrago-Garcia, D. C. et al. Asymptomatic SARS-CoV-2 infections: a living systematic review and meta-analysis. medRxiv 2020.04.25.20079103 (2020).

22. Wang, X. et al. Impact of Social Distancing Measures on Coronavirus Disease Healthcare Demand, Central Texas, USA. Emerg. Infect. Dis. 26, 2361–2369 (2020).

23. Letizia, A. G. et al. SARS-CoV-2 Transmission among Marine Recruits during Quarantine. N. Engl. J. Med. (2020) doi:10.1056/NEJMoa2029717.

24. Michael, N. L. SARS-CoV-2 in the U.S. Military - Lessons for Civil Society. N. Engl. J. Med. (2020) doi:10.1056/NEJMe2032179.

25. Matson, Z. College restart nears as students are asked to quarantine. The Daily Gazette (2020).

26. COVID-19 testing strategies vary widely across institutions. https://www.insidehighered.com/news/2020/08/21/covid-19-testing-strategies-vary-widely-across-institutions.

27. Sethuraman, N., Jeremiah, S. S. & Ryo, A. Interpreting Diagnostic Tests for SARS-CoV-2. JAMA 323, 2249–2251 (2020).

28. Wang, Y. et al. Characterization of an Asymptomatic Cohort of Severe Acute Respiratory Syndrome Coronavirus 2 (SARS-CoV-2) Infected Individuals Outside of Wuhan, China. Clinical Infectious Diseases (2020) doi:10.1093/cid/ciaa629.

29. Li, Y. et al. Asymptomatic and Symptomatic Patients With Non-severe Coronavirus Disease (COVID-19) Have Similar Clinical Features and Virological Courses: A Retrospective Single Center Study. Front. Microbiol. 11, 1570 (2020).

30. Lee, S. et al. Clinical Course and Molecular Viral Shedding Among Asymptomatic and Symptomatic Patients With SARS-CoV-2 Infection in a Community Treatment Center in the Republic of Korea. JAMA Intern. Med. (2020) doi:10.1001/jamainternmed.2020.3862.

31. Ferretti, L. et al. The timing of COVID-19 transmission. medRxiv 2020.09.04.20188516 (2020).

32. CDC. Duration of Isolation and Precautions for Adults with COVID-19. https://www.cdc.gov/coronavirus/2019-ncov/hcp/duration-isolation.html (2020).

33. Xiao, A. T. et al. Dynamic profile of RT-PCR findings from 301 COVID-19 patients in Wuhan, China: A descriptive study. J. Clin. Virol. 127, 104346 (2020).

34. Cevik M, Tate M, Lloyd O, Maraolo AE, Schafers J, Ho A. SARS-CoV-2, SARS-CoV, and MERS-CoV viral load dynamics, duration of viral shedding, and infectiousness: a systematic review and meta-analysis. The Lancet Microbe (2020) doi:10.1016/S2666-5247(20)30172-5.

35. Dinnes, J. et al. Rapid, point-of-care antigen and molecular-based tests for diagnosis of SARS-CoV-2 infection. Cochrane Database Syst. Rev. 8, CD013705 (2020).

36. Zhang, Y., Li, Y., Wang, L., Li, M. & Zhou, X. Evaluating Transmission Heterogeneity and Super-Spreading Event of COVID-19 in a Metropolis of China. Int. J. Environ. Res. Public Health 17, (2020).

37. Endo, A., Abbott, S., Kucharski, A. J., Funk, S. & Centre for the Mathematical Modelling of Infectious Diseases COVID-19 Working Group. Estimating the overdispersion in COVID-19 transmission using outbreak sizes outside China. Wellcome Open Research vol. 5 67 (2020).

38. Website.. https://assets.researchsquare.com/files/rs-29548/v1_stamped.pdf.

39. Bi, Q. et al. Epidemiology and transmission of COVID-19 in 391 cases and 1286 of their close contacts in Shenzhen, China: a retrospective cohort study. The Lancet Infectious Diseases vol. 20 911–919 (2020).

## References

1. Li, Y. et al. Asymptomatic and Symptomatic Patients With Non-severe Coronavirus Disease (COVID-19) Have Similar Clinical Features and Virological Courses: A Retrospective Single Center Study. Front. Microbiol. 11, 1570 (2020).

2. Lee, S. et al. Clinical Course and Molecular Viral Shedding Among Asymptomatic and Symptomatic Patients With SARS-CoV-2 Infection in a Community Treatment Center in the Republic of Korea. JAMA Intern. Med. (2020) doi:10.1001/jamainternmed.2020.3862.

3. Zhang, Y., Li, Y., Wang, L., Li, M. & Zhou, X. Evaluating Transmission Heterogeneity and Super-Spreading Event of COVID-19 in a Metropolis of China. Int. J. Environ. Res. Public Health 17, (2020).

4. He, X. et al. Author Correction: Temporal dynamics in viral shedding and transmissibility of COVID-19. Nat. Med. (2020) doi:10.1038/s41591-020-1016-z.

5. Meeker, W. Q. & Escobar, L. A. Teaching about Approximate Confidence Regions Based on Maximum Likelihood Estimation. The American Statistician vol. 49 48 (1995).

6. Miller, T. E. et al. Clinical sensitivity and interpretation of PCR and serological COVID-19 diagnostics for patients presenting to the hospital. FASEB J. (2020) doi:10.1096/fj.202001700RR.

7. Moghadas, S. M. et al. Projecting hospital utilization during the COVID-19 outbreaks in the United States. Proc. Natl. Acad. Sci. U. S. A. 117, 9122–9126 (2020).

8. Qin, J. et al. Estimation of incubation period distribution of COVID-19 using disease onset forward time: A novel cross-sectional and forward follow-up study. Sci Adv 6, eabc1202 (2020).

9. Xiao, A. T. et al. Dynamic profile of RT-PCR findings from 301 COVID-19 patients in Wuhan, China: A descriptive study. J. Clin. Virol. 127, 104346 (2020).

10. CDC. Duration of Isolation and Precautions for Adults with COVID-19. https://www.cdc.gov/coronavirus/2019-ncov/hcp/duration-isolation.html (2020).

11. Cevik M, Tate M, Lloyd O, Maraolo AE, Schafers J, Ho A. SARS-CoV-2, SARS-CoV, and MERS-CoV viral load dynamics, duration of viral shedding, and infectiousness: a systematic review and meta-analysis. The Lancet Microbe (2020) doi:10.1016/S2666-5247(20)30172-5.

12. Nishiura, H. et al. Estimation of the asymptomatic ratio of novel coronavirus infections (COVID-19). Int. J. Infect. Dis. 94, 154–155 (2020).

13. Buitrago-Garcia, D. C. et al. Asymptomatic SARS-CoV-2 infections: a living systematic review and meta-analysis. medRxiv 2020.04.25.20079103 (2020).

14. Wang, Y. et al. Characterization of an Asymptomatic Cohort of Severe Acute Respiratory Syndrome Coronavirus 2 (SARS-CoV-2) Infected Individuals Outside of Wuhan, China. Clinical Infectious Diseases (2020) doi:10.1093/cid/ciaa629.

15. Wang, X. et al. Impact of Social Distancing Measures on Coronavirus Disease Healthcare Demand, Central Texas, USA. Emerg. Infect. Dis. 26, 2361–2369 (2020).

